# Microbiota compositions from infertile couples seeking *in vitro* fertilization (IVF), using 16S rRNA gene sequencing methods: any correlation to clinical outcomes?

**DOI:** 10.1101/2020.10.22.20215988

**Authors:** Somadina I Okwelogu, Joseph I Ikechebelu, Nneka R Agbakoba, Kingsley C Anukam

## Abstract

Bacterial infections are usually suspected in infertile couples seeking IVF with no clear understanding of the microbial compositions present in the seminal fluids and vaginal swabs of the patients. We used next-generation sequencing technology to correlate microbiota compositions with IVF clinical outcomes. Thirty-six couples were recruited to provide seminal fluids and vaginal swabs. Seminal fluid microbiota compositions had lower bacterial concentrations compared with the vagina, but species diversity was significantly higher in seminal fluid samples. Azoospermic subjects had more relative abundance of *Mycoplasma* and *Ureaplasma*. In Normospermic semen *Lactobacillus* (43.86%) was the most abundant, followed by *Gardnerella* (25.45%), while the corresponding vaginal samples, *Lactobacillus* (61.74%) was the most abundant, followed by *Prevotella* (6.07%), and *Gardnerella* (5.86%). Semen samples with positive IVF were significantly colonized by *Lactobacillus jensenii* (*P*=0.002), *Faecalibacterium* (*P*=0.042) and significantly less colonized by *Proteobacteria, Prevotella, Bacteroides* and lower *Firmicutes*/*Bacteroidetes* ratio compared with semen samples with negative IVF. Vaginal samples with positive IVF clinical outcome were significantly colonized by *Lactobacillus gasseri*, less colonized by *Bacteroides*, and *Lactobacillus iners*. This study has opened a window of possibility for *Lactobacillus* replenishments in men and women prior to IVF treatment.

## Introduction

Bacterial infections affecting the reproductive tracts of males and or females with infertility have been documented in previous studies using culture methods ^**^1, 2^**^. Some bacteria, fungi, viruses and parasites are known to interfere with reproductive functions in both male and females of reproductive age and infections of the genitourinary tract account for about 15% of male infertility cases. More than a few bacteria, including *Lactobacillus iners, Gardnerella vaginalis, Escherichia faecalis, E. coli* and *Staphylococcus aureus*, have been found to be associated with male infertility as demonstrated using polymerase chain reaction (PCR)^**3**^. Bacterial vaginosis (BV) has been found to be a major risk factor for infertility^4^. Some specific bacteria incriminated in BV such as *Atopobium vaginae, Ureaplasma vaginae, U. parvum, U. urealyticum, Gardnerella vaginalis* and reduced abundance of *Lactobacillus* species are associated with infertility in women^5^. Women with endometrial dysbiosis have also been found to experience implantation failure leading to infertility^**6**^. While vaginal microbiota is normally under the influence of oestrogen^**7**^, endometrial microbiota is not affected by hormonal fluctuations^**8**^

There are several studies confirming that a vaginal microbiota replete with relative abundances of Lactobacillus species, devoid of bacterial vaginosis, leads to more positive IVF clinical outcomes^**9**^

Besides other sexually-transmitted infections, genital mycoplasmas are associated with poor reproductive health of women including but not limited to endometritis, cervicitis and pelvic inflammatory disease and adverse pregnancy outcomes^**10**,**11**^. The pregnancy success rate of the various assisted reproductive health care such as in vitro fertilization (IVF) tend to be reduced as a result of prior *mycoplasma* colonization of the female and male genital tract^**12**^. One wonders whether genital *Mycoplasma* was the only pathogen associated with poor pregnancy outcome, although several studies have shown that *Mycoplasma* species can attach to spermatozoa and remain adherent to spermatozoa after assisted reproductive treatment washing procedures^**13**,**14**^.

Clinical studies have shown that bacterial contamination of the embryo transfer catheter has significant negative effect on the clinical pregnancy rates^**15**^. Approximately, 35% of infertile women are afflicted with post-inflammatory changes of the oviduct or surrounding peritoneum that interfere with tubal-ovarian function mostly as a result of infection and are likely to develop ectopic (tubal) pregnancy^**16**^.

In Africa, especially Nigeria, little is known about the bacterial communities found in the seminal fluids of men seeking reproductive health care with next-generation sequencing technology. A recent pilot study of seminal fluid in a tertiary hospital revealed varying bacterial community diversities that are unique in each sample in contrast to culture-dependent methods^**17**^. We have also previously shown that women with bacterial vaginosis (BV) had varying proportions of diverse bacteria including *Lactobacillus* species in all BV subjects, but the total number of all the BV-associated microbes (*Gardnerella, Prevotella, Magasphaera*, and others) outnumbered *Lactobacillus* genera^**18**^. In the present study, next-generation 16S rRNA gene sequencing method was used to compare seminal bacterial composition in couples seeking reproductive health care-IVF. In addition, the study delineated semen quality, bacterial functional gene predictions and correlated microbiota composition with clinical outcome of the IVF assisted reproductive care.

## Results

### Demographic information, IVF clinical outcome and semen quality

As shown in **Table 1a/Table 1b**, of the 36 men that were examined for semen quality and that had result for 16S rRNA gene sequencing, 11 were clinically diagnosed as having primary infertility with duration of infertility ranging from 1-13 years, while 25 men were diagnosed with secondary infertility, had duration of infertility from 1-8 years. The semen characteristics of the subjects were as follows; 11 samples were assigned as normospermia (>15×10^6^), 7 had oligospermia (<15×10^6^), 7 had azoospermia, 10 had asthenozoospermia, while 1 sample was classified as tetratozoospermia. The number of semen samples that had leucocytes (pyospermia) were 11, while 1 sample was assigned as oligoasthenozoospermia.

**Table 1a:** Demographic information and IVF clinical outcome

| **Sample No.** | **Female age range (years)** | **Clinical Pregnancy outcome** | **Scan result** | **Sex of the baby** | **Live delivery** | **Male age range (years)** | **Duration of marriage (years)** | **Duration of infertility (years)** | **Type of infertility** |
| --- | --- | --- | --- | --- | --- | --- | --- | --- | --- |
| 1 | 25-29 | Negative | - | - |  | 35-39 | 5-9 | 5-9 | Primary |
| 2 | 35-39 | Negative | - | - |  | 40-44 | 10-14 | 5-9 | Secondary |
| 3 | 40-44 | Negative | - | - |  | 45-49 | 5-9 | 5-9 | Primary |
| 4 | 30-34 | Negative | - | - |  | 35-39 | 10-14 | 10-14 | Primary |
| 5 | 30-34 | Negative | - | - |  | 35-39 | 5-9 | 5-9 | Primary |
| 6 | 25-29 | Negative | - | - |  | 25-29 | 0-4 | 0-4 | Primary |
| 7 | 35-39 | POSITIVE | Singleton | Male | YES | 45-49 | 5-9 | 5-9 | Secondary |
| 8 | 30-34 | Negative | - | - |  | 30-34 | 0-4 | 0-4 | Primary |
| 9 | 25-29 | Negative | - | - |  | 25-29 | 0-4 | 0-4 | Primary |
| 10 | 35-39 | POSITIVE | Singleton | Female | YES | 40-44 | 5-9 | 5-9 | Primary |
| 11 | 40-44 | POSITIVE | Twins | Male, Female | YES | 45-49 | 0-4 | 0-4 | Secondary |
| 12 | 40-44 | Negative | - | - |  | 40-44 | 0-4 | 0-4 | Primary |
| 13 | 23-29 | Negative | - | - |  | 30-34 | 0-4 | 0-4 | Primary |
| 14 | 30-34 | POSITIVE | Singleton | Male | YES | 35-39 | 0-4 | 0-4 | Primary |
| 15 | 25-29 | Negative | - | - |  | 25-29 | 5-9 | 0-4 | Secondary |
| 16 | 30-34 | POSITIVE | Singleton | Male | YES | 30-34 | 0-4 | 0-4 | Primary |
| 17 | 30-34 | POSITIVE | Singleton | Male | YES | 35-39 | 0-4 | 0-4 | Primary |
| 18 | 40-44 | Negative | - | - |  | 50-54 | 5-9 | 5-9 | Secondary |
| 19 | 30-34 | POSITIVE | Twins | Male, Female | YES | 40-44 | 0-4 | 0-4 | Primary |
| 20 | 30-34 | Negative | - | - |  | 30-34 | 5-9 | 5-9 | Primary |
| 21 | 35-39 | Negative | - | - |  | 45-49 | 5-9 | 5-9 | Secondary |
| 22 | 30-34 | Negative | - | - |  | 40-44 | 0-4 | 0-4 | Secondary |
| 23 | 40-44 | POSITIVE | Twins | Male, Female | YES | 50-54 | 10-14 | 5-9 | Secondary |
| 24 | 30-34 | Negative | - | - |  | 30-34 | 5-9 | 5-9 | Primary |
| 25 | 25-29 | POSITIVE | Singleton | Female | No<br>(Misca) | 30-34 | 0-4 | 0-4 | Primary |
| 26 | 45-49 | Negative | - | - |  | 50-54 | 0-4 | 0-4 | Primary |
| 27 | 30-34 | POSITIVE | Singleton | Male | YES | 35-39 | 5-9 | 5-9 | Primary |
| 28 | 30-34 | Negative | - | - |  | 35-39 | 0-4 | 0-4 | Primary |
| 29 | 30-34 | Negative | - | - |  | 30-34 | 0-4 | 0-4 | Primary |
| 30 | 30-34 | Negative | - | - |  | 35-39 | 5-9 | 0-4 | Secondary |
| 31 | 45-49 | Negative | - | - |  | 60-64 | 10-14 | 10-14 | Primary |
| 32 | 40-44 | Negative | - | - |  | 45-49 | 5-9 | 5-9 | Primary |
| 33 | 40-44 | POSITIVE | Singleton | Male | YES | 50-54 | 5-9 | 5-9 | Primary |
| 34 | 35-39 | Negative | - | - |  | 50-54 | 10-14 | 5-9 | Secondary |
| 35 | 30-34 | POSITIVE | Singleton | Female | YES | 40-44 | 0-4 | 0-4 | Secondary |
| 36 | 33-39 | Negative | - | - |  | 35-39 | 5-9 | 5-9 | Primary |

**Table 1b:** Semen Characteristics

| Sample # | Sexual Intercourse abstinence (days) | Viscosity | Liquefaction | Volume (ml) | Progressive motility (%) | Non-Progressive motility (%) | Non-motile (%) | Total motility (%) | Velocity (mic/sec) | Sperm conc (m/ml) | Total sperm/ Volume (million) | Presence of pus cells |
| --- | --- | --- | --- | --- | --- | --- | --- | --- | --- | --- | --- | --- |
| 1 | 3 | NORMAL | NORMAL | 2 | 9 | 3 | 88 | 12 | 32 | 88 | 176.1 | + |
| 2 | 5 | NORMAL | NORMAL | 2.5 | 5 | 2 | 93 | 7 | 27 | 22.2 | 55.4 | NIL |
| 3 | 4 | ABNORMAL | ABNORMAL | 1.2 | 0 | 0 | 0 | 0 | 0 | 0 | 0 | 2+ |
| 4 | 3 | NORMAL | NORMAL | 2.1 | 2 | 9 | 89 | 11 | 0 | 2 | 0 | NIL |
| 5 | 3 | NORMAL | NORMAL | 1.2 | 27 | 6 | 67 | 33 | 41 | 120.5 | 144.6 | NIL |
| 6 | 4 | ABNORMAL | ABNORMAL | 1.2 | 0 | 0 | 0 | 0 | 0 | 0 | 0 | 2+ |
| 7 | 3 | NORMAL | NORMAL | 2.8 | 37 | 10 | 53 | 47 | 38 | 77.4 | 216.7 | NIL |
| 8 | 3 | NORMAL | NORMAL | 1 | 44 | 8 | 48 | 52 | 43 | 72 | 72 | NIL |
| 9 | 4 | NORMAL | NORMAL | 3 | 22 | 10 | 68 | 32 | 30 | 33.8 | 101.4 | NIL |
| 10 | 5 | NORMAL | NORMAL | 1.5 | 39 | 8 | 53 | 47 | 42 | 194.5 | 291.8 | NIL |
| 11 | 3 | NORMAL | NORMAL | 2.3 | 51 | 12 | 37 | 63 | 39 | 52.7 | 121.3 | NIL |
| 12 | 5 | NORMAL | NORMAL | 0.8 | 17 | 3 | 80 | 20 | 46 | 207.6 | 166.1 | NIL |
| 13 | 4 | NORMAL | NORMAL | 2 | 52 | 20 | 28 | 72 | 32 | 6.4 | 12.8 | NIL |
| 14 | 4 | ABNORMAL | ABNORMAL | 3 | 0 | 0 | 0 | 0 | 0 | 0 | 0 | 3+ |
| 15 | 3 | NORMAL | NORMAL | 3.2 | 0 | 1 | 99 | 1 | 0 | 16 | 51.1 | NIL |
| 16 | 5 | NORMAL | NORMAL | 2.2 | 35 | 8 | 57 | 43 | 42 | 77.2 | 169.8 | NIL |
| 17 | 3 | NORMAL | NORMAL | 1.5 | 11 | 7 | 82 | 18 | 25 | 66.4 | 99.7 | NIL |
| 18 | 3 | ABNORMAL | ABNORMAL | 2.5 | 0 | 0 | 0 | 0 | 0 | 0 | 0 | 2+ |
| 19 | 4 | NORMAL | NORMAL | 1.5 | 30 | 6 | 62 | 38 | 43 | 163.3 | 244.9 | NIL |
| 20 | 3 | ABNORMAL | ABNORMAL | 5 | 0 | 0 | 0 | 0 | 0 | 0 | 0 | 2+ |
| 21 | 4 | NORMAL | NORMAL | 3.2 | 46 | 9 | 40 | 60 | 44 | 50.3 | 160.9 | NIL |
| 22 | 5 | ABNORMAL | ABNORMAL | 2 | 0 | 0 | 0 | 0 | 0 | 0 | 0 | 2+ |
| 23 | 3 | NORMAL | NORMAL | 2 | 50 | 4 | 84 | 16 | 34 | 157 | 314 | 2+ |
| 24 | 3 | NORMAL | NORMAL | 3.2 | 47 | 10 | 40 | 60 | 45 | 61.6 | 197.2 | NIL |
| 25 | 3 | ABNORMAL | ABNORMAL | 2.1 | 0 | 0 | 0 | 0 | 0 | 0 | 0 | 2+ |
| 26 | 4 | NORMAL | NORMAL | 2 | 5 | 24 | 29 | 29 | 5 | 4.7 | 9.4 | NIL |
| 27 | 3 | NORMAL | NORMAL | 2.4 | 3 | 2 | 95 | 5 | 22 | 75.3 | 180 | NIL |
| 28 | 4 | NORMAL | NORMAL | 2 | 40 | 14 | 46 | 54 | 34 | 44.6 | 89.2 | NIL |
| 29 | 3 | NORMAL | NORMAL | 1 | 3 | 2 | 95 | 5 | 22 | 72.2 | 72.2 | NIL |
| 30 | 5 | NORMAL | NORMAL | 3 | 8 | 19 | 73 | 27 | 10 | 5.6 | 16.9 | 3+ |
| 31 | 3 | NORMAL | NORMAL | 3.2 | 1 | 11 | 88 | 12 | 4 | 11.3 | 36 | NIL |
| 32 | 4 | ABNORMAL | ABNORMAL | 2.1 | 39 | 22 | 39 | 61 | 26 | 5.1 | 10.8 | 2+ |
| 33 | 5 | NORMAL | NORMAL | 1.5 | 27 | 6 | 67 | 33 | 40 | 77.6 | 116.4 | NIL |
| 34 | 4 | NORMAL | NORMAL | 1 | 20 | 5 | 75 | 25 | 37 | 134.1 | 134.1 | NIL |
| 35 | 3 | NORMAL | NORMAL | 2.5 | 29 | 9 | 62 | 38 | 34 | 80.8 | 97 | NIL |
| 36 | 5 | NORMAL | NORMAL | 3 | 44 | 7 | 49 | 51 | 45 | 83.8 | 251.5 | NIL |

### Semen and Vagina microbiome compositions

The first objective was to determine whether seminal fluid microbiota differ substantially with the vagina. In this regard, the alpha diversity that estimates the species richness typified by ACE, CHAO Jackknife, Shannon and Simpson, shows that seminal fluid microbiota composition is less in species richness (lower bacterial concentrations) compared with the vagina as shown in **Fig. 1a**.

**Fig. 1a/1b:**
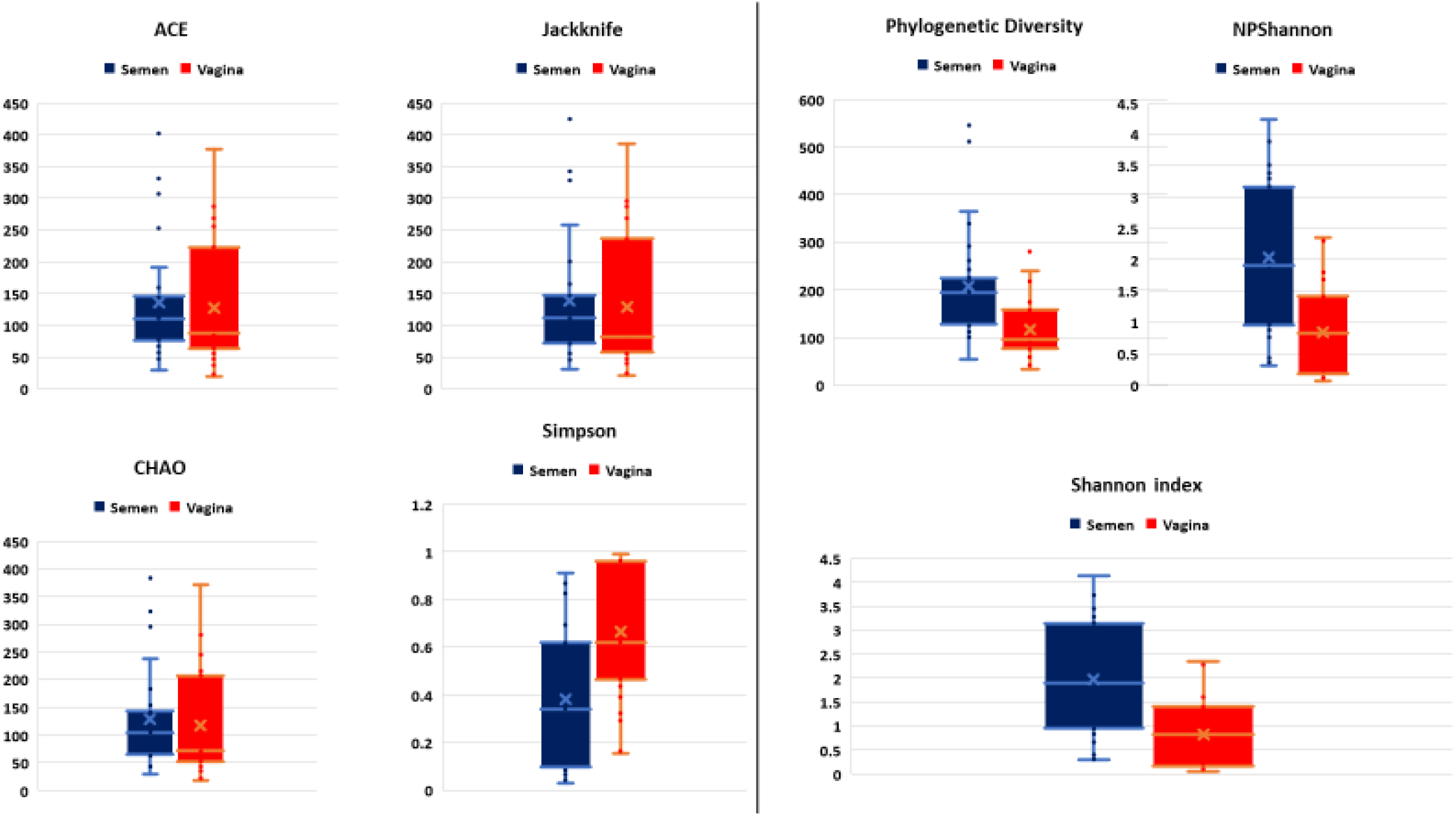
Comparative alpha diversities between semen and vagina showing species richness typified by ACE, CHAO Jackknife, Shannon and Simpson index.

However, species diversity was significantly higher in seminal fluid samples represented in **Fig. 1b**. The Principle Co-Ordinate Analysis (PCoA) with Bray-curtis metrices confirmed the differences in bacterial community diversities between semen and vagina as shown in Supplementary **Fig. S1**. The EzBiocloud Microbiome Taxonomic Profile (MTP) pipeline was able to provide distinct taxonomic categories (Family, Genus and Species) at 1% cut-off between semen and vagina as shown in **Fig. 2**.

**Fig. 2:**
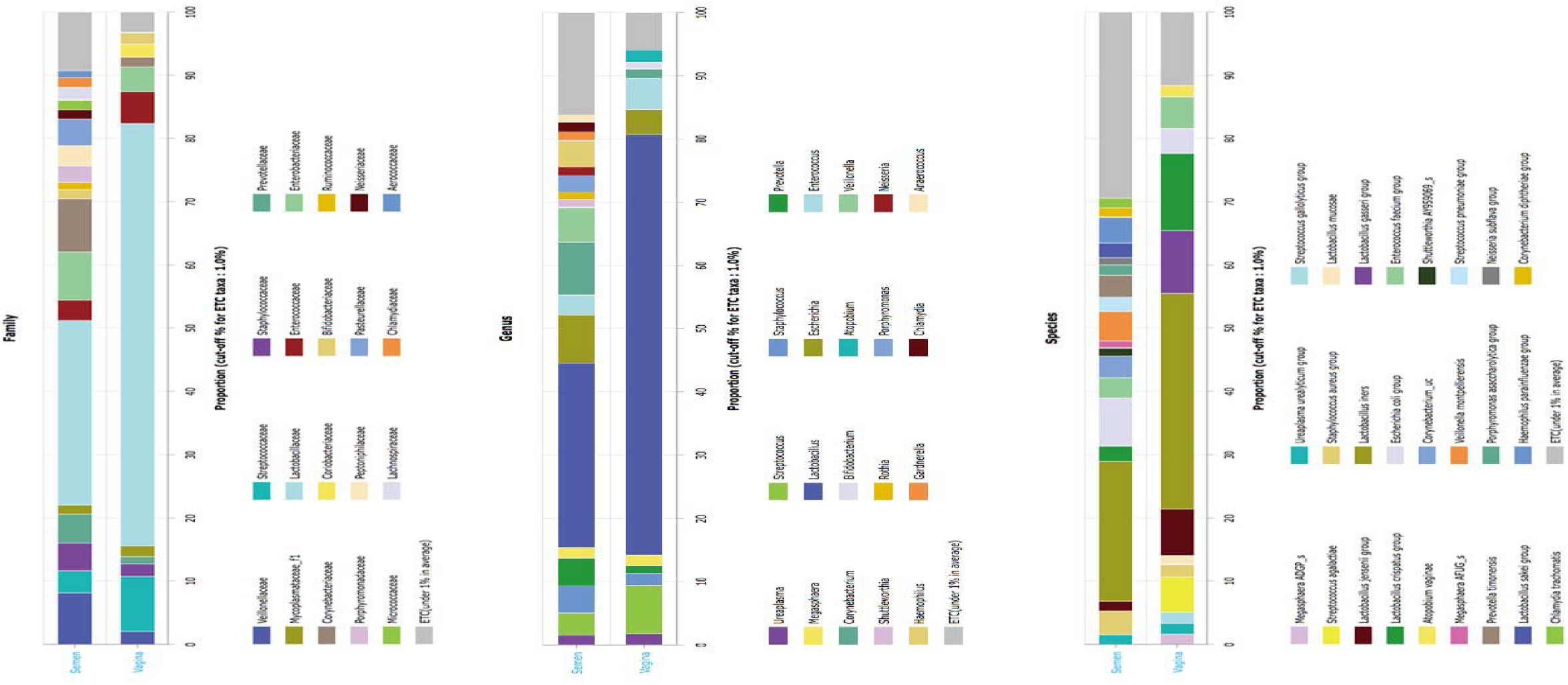
EzBiocloud Microbiome Taxonomic Profile (MTP) pipeline showing distinct taxonomic categories (Family, Genus and Species) at 1% cut-off between semen and vagina.

The rarefaction curve showing the number of reads between the semen samples and vaginal microbiota is presented in Supplementary **Fig. S2**. For comparative purposes, we selected the corresponding semen and vagina bacterial communities in line with the three semen categories. At the genera taxonomic level, out of 621 genera identified, 6.8% were exclusive to normospermia, 20.5% to oligospermia and 2.9% exclusive to azoospermia, while 41.5% were common to all the categories. When the genera taxa are examined with the three semen categories, *Mycoplasma* and *Ureaplasma* occurred more in relative abundance in azoospermic subjects (**Fig. 3**).

**Fig. 3:**
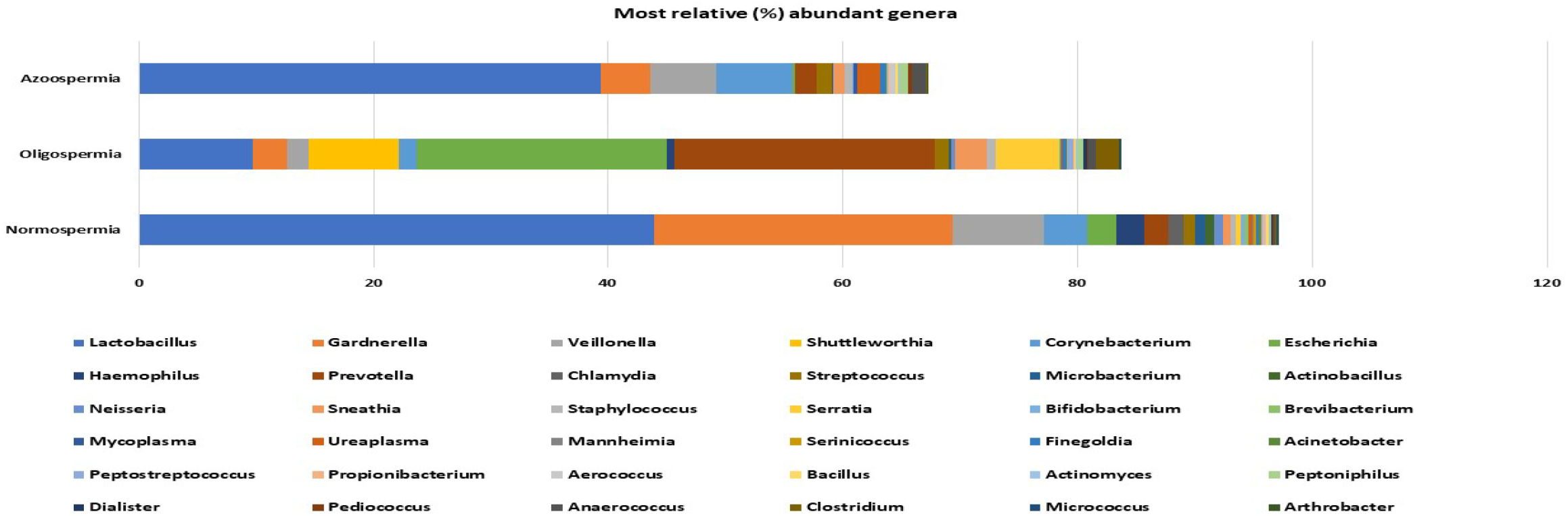
Most relative (%) genera abundance between the semen categories.

At the species taxonomic level, out of 1384 species identified, 10.3% were exclusive to normospermia, 26.4% to oligospermia and 6.8% exclusive to azoospermia, while 27% were common to all the categories as shown in Supplementary **Fig. S3**.

### Microbiota compositions from Normospermia couples

Twenty-four phyla were identified from subjects with normospermia, while 22 were identified from the corresponding vagina samples. *Firmicutes* accounted for 54.47% vs 75.89% relative abundance, followed by *Actinobacteria* (32.26% vs 8.72%), *Proteobacteria* (8.60% vs 7.20%), *Bacteroidetes* (2.29% vs 5.95%), *Chlamydiae* (1.32% vs 0.0003%), *Fusobacteria* (0.67% vs 2.09%), *Tenericutes* (0.30% vs 0.09%) and others represented in Supplementary **Fig. S4**. At the genera taxonomic level, 451 genera were identified in normospermia samples while 331 genera were found in the corresponding females. *Lactobacillus* (43.86%) was the most abundant genera in semen, followed by *Gardnerella* (25.45%), *Veillonella* (7.78%), *Corynebacterium* (3.73%), *Escherichia* (2.47%), *Haemophilus* (2.36%), *Prevotella* (2.03%) and others, while in the corresponding vagina samples, *Lactobacillus* (61.74%) was the most abundant genera, followed by *Prevotella* (6.07%), *Gardnerella* (5.86%), *Streptococcus* (5.84%), *Escherichia* (5.40%), *Megasphaera* (4.51%), *Sneathia* (2.13%) and others represented in Supplementary **Fig. S5**.

At the species taxonomic level, 848 species were identified in normospermia samples, while 585 species were found in the corresponding vagina samples. *Gardnerella vaginalis* (31.93%) was the most abundant species identified in 10/12 of the semen samples, followed by *Lactobacillus iners* 8/12 (15.56%), *Lactobacillus pentosus* 1/12 (12.39%), *Veillonella montpellierensis* 8/12 (9.61%), *Lactobacillus japonicus* 3/12 (4.29%), *Haemophilus parainfluenzae* 9/12 (2.91%), *Corynebacterium tuberculostearicum* 11/12 (1.75%), *Lactobacillus jensenii* 3/12 (1.73%) and others. The corresponding vaginal samples, had *Lactobacillus iners* 9/12 (49.06%), as the most abundant species, followed by *Lactobacillus jensenii* 2/12 (10.60%), *Peptostreptococcus stomatis* 6/12 (6.92%), *Actinocatenispora silicis* 5/12 (6.19%), *Pasteurella pneumotropica* 2/12 (4.09%), *Actinomyces naturae* 4/12 (3.32%), *Lactobacillus taiwanensis* 10/12 (2.78%) and *Peptoniphilus asaccharolyticus* 8/12 (2.69%) and others as shown in **Fig. 4**.

**Fig. 4:**
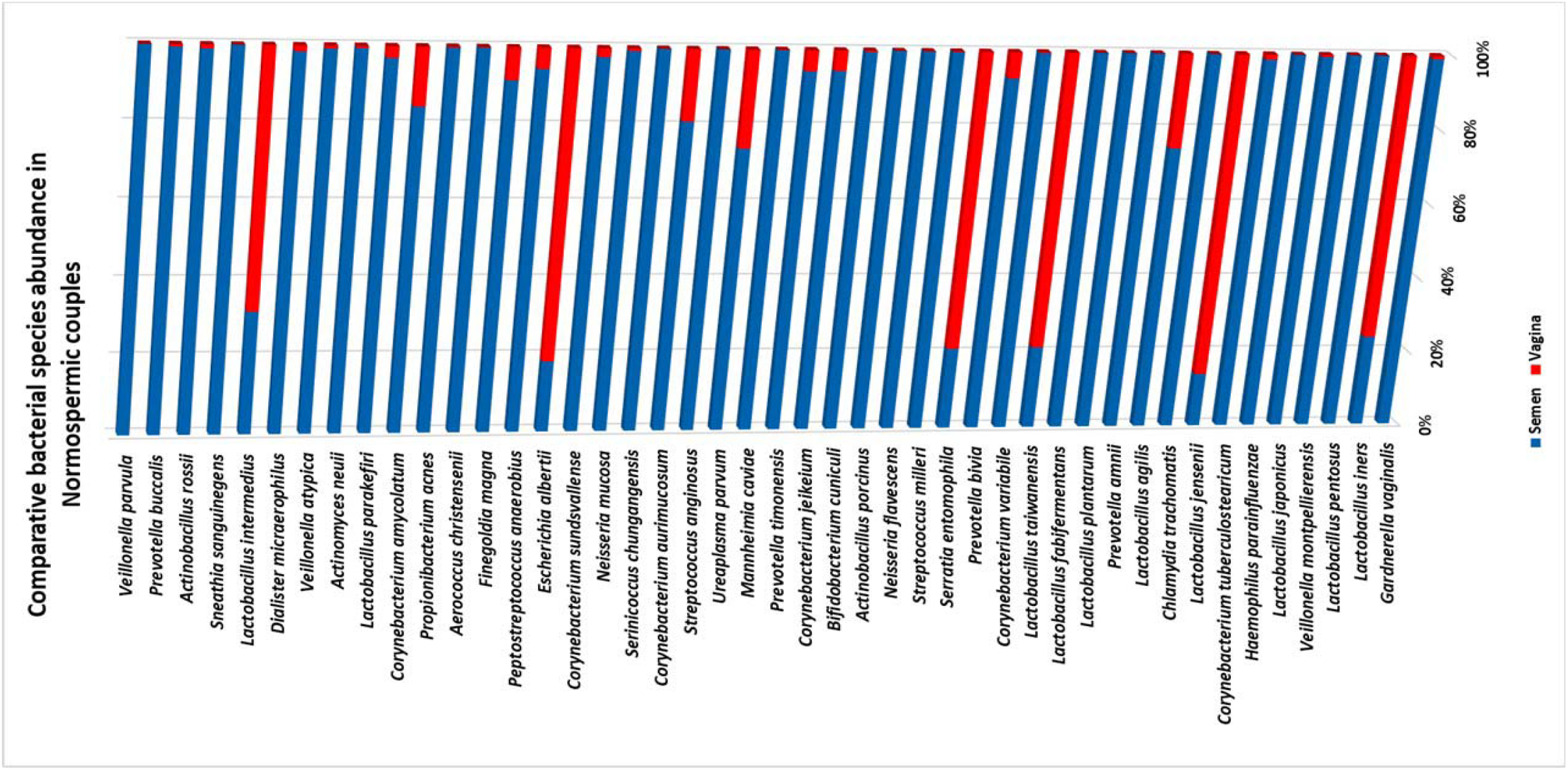
Comparative relative (%) species abundance in normospermia couples.

### Microbiota compositions from oligospermia couples

Among the oligospermic couples, 555 genera were identified in semen samples, while 403 genera were found in the corresponding vaginal samples. *Prevotella* (22.13%) was the most abundant genus in oligospermic semen, followed by *Escherichia* (21.33%), *Lactobacillus* (9.73%), *Shuttleworthia* (7.67%), *Serratia* (5.33%), *Megasphaera* (5.04%), *Gardnerella* (2.85%), *Sneathia* (2.79%), *Porphyromonas* (2.22%) and others. The corresponding vaginal samples had *Lactobacillus* (53.90%) as the most abundant genus, followed by *Streptococcus* (14.68%), *Gardnerella* (5.48%) and others as shown in **Fig. 5a**.

**Fig. 5a,5b:**
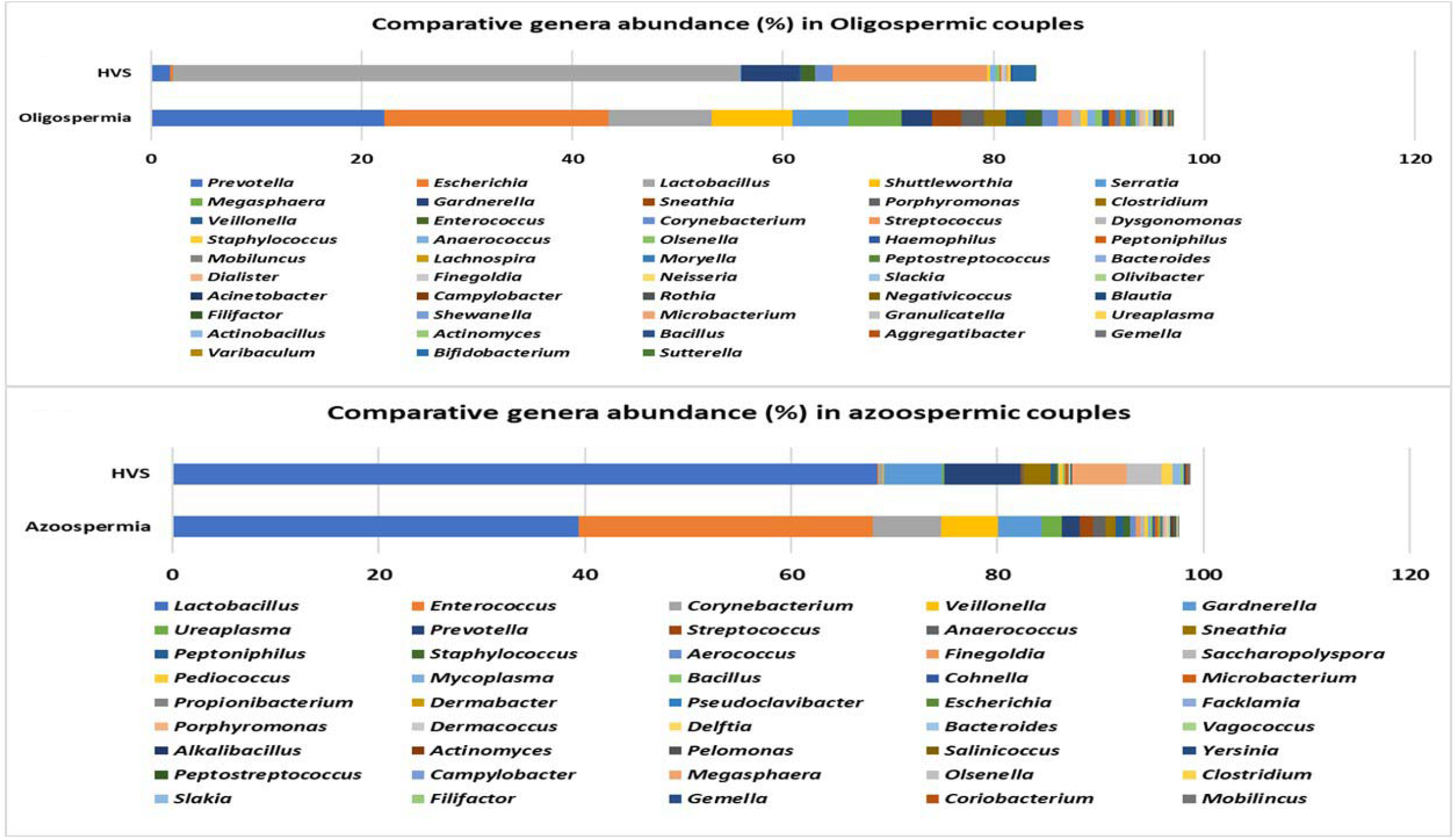
Comparative relative (%) genera abundance in oligospermia and azoospermia couples.

**Fig. 6:**
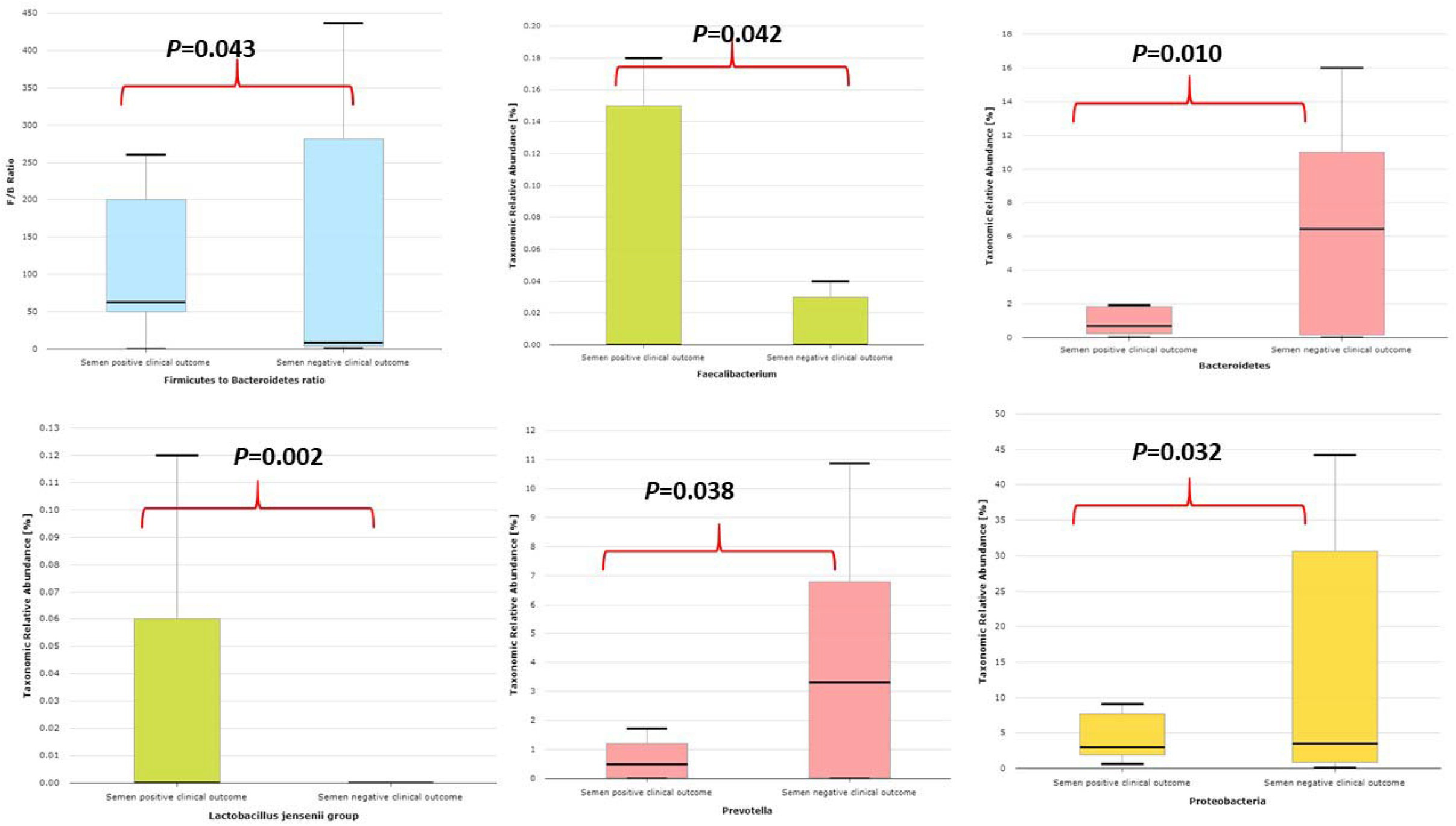
Comparative relative abundance of some selected taxa showing significant difference between semen samples with positive IVF clinical outcome and samples with negative IVF clinical outcome.

**Fig. 7:**
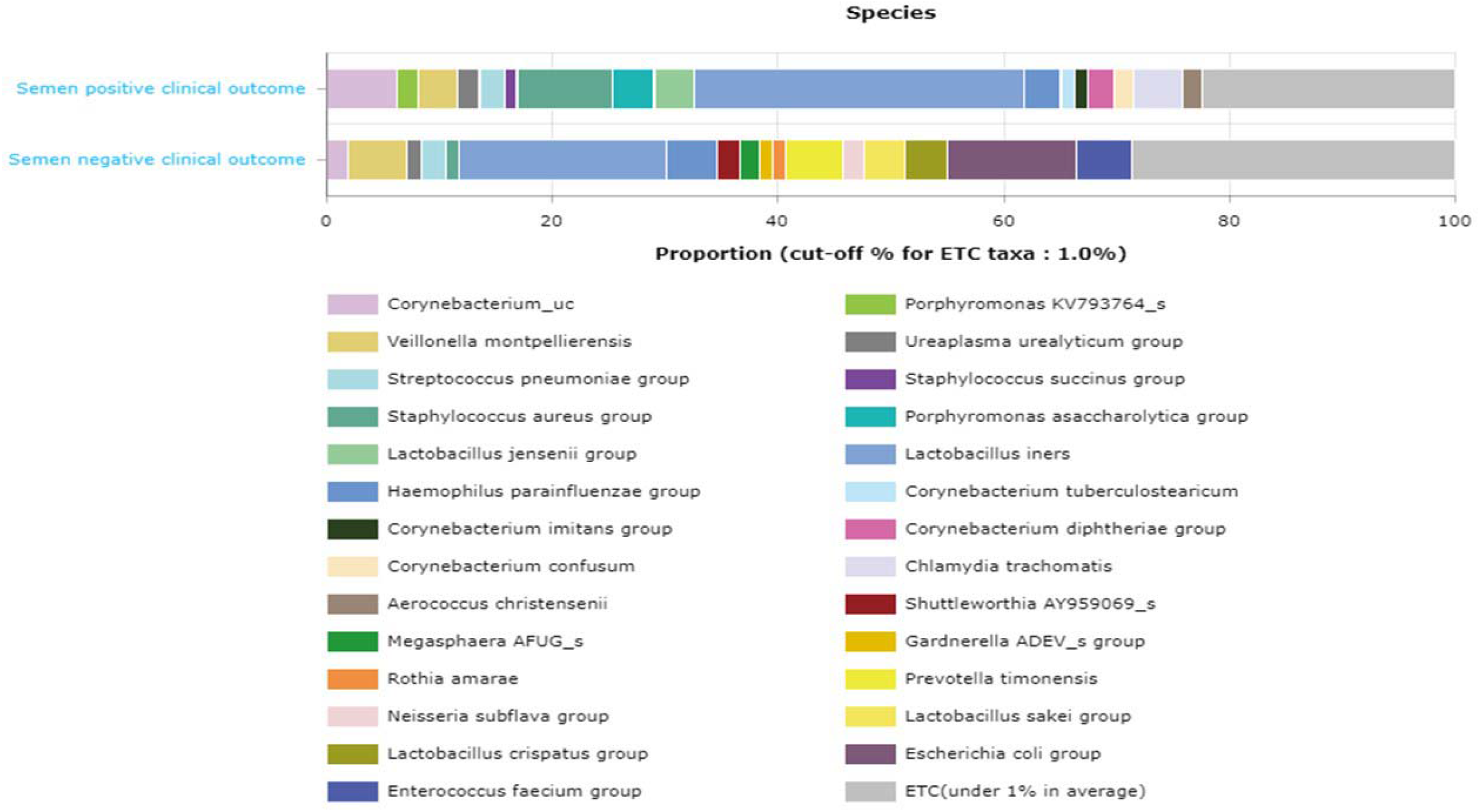
Comparative proportion (cut-off 1.0%) of the relative abundance at the species taxonomic level between semen samples with positive IVF clinical outcome and samples with negative IVF clinical outcome.

**Fig. 8:**
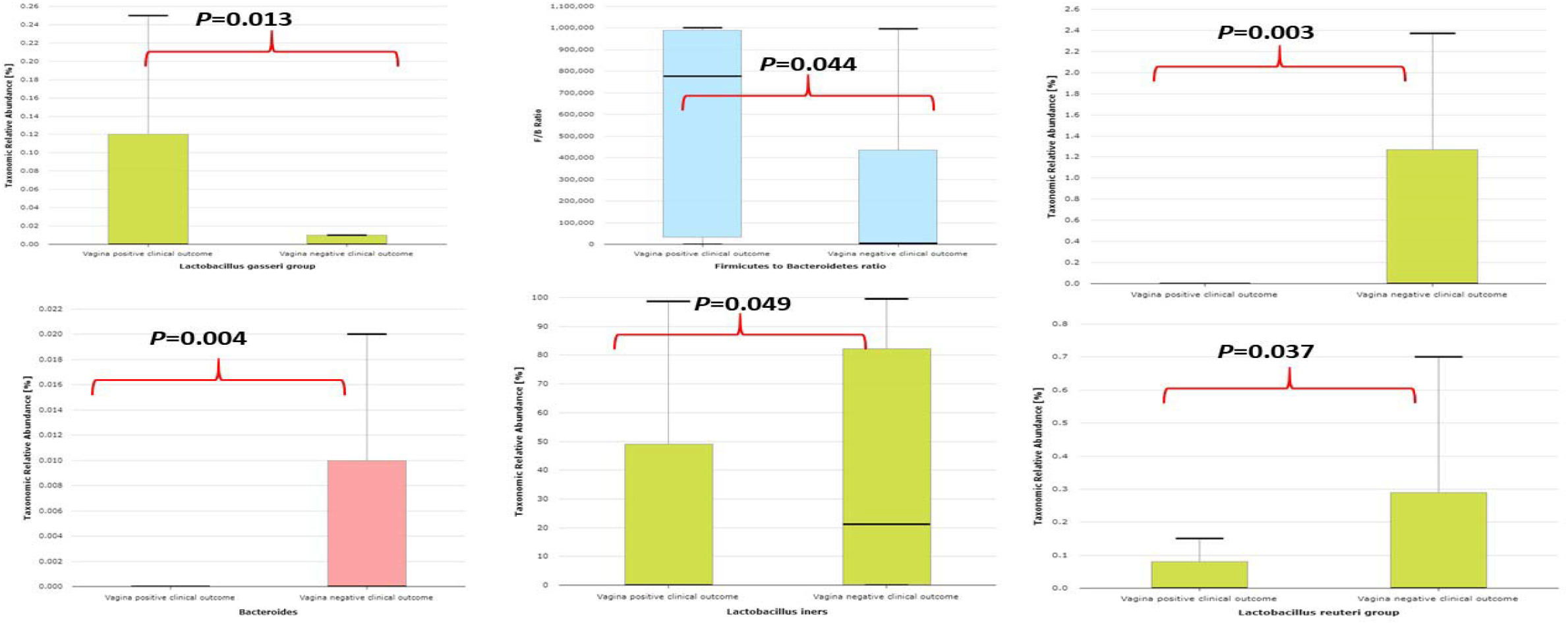
Comparative relative abundance of some selected taxa showing significant difference between vaginal samples with positive IVF clinical outcome and vaginal samples with negative IVF clinical outcome.

### Microbiota compositions from azoospermia couples

The results from azoospermic couples shows that 342 genera colonized the semen samples while 309 genera were found in the corresponding vaginal samples. Lactobacillus (39.38%) was the most abundant genus in azoospermic men, followed by Enterococcus (28.52%), Corynebacterium (6.58%), Veillonella (5.53%), Gardnerella (4.25%), Ureaplasma (1.91%), Prevotella (1.82%) and others. The corresponding vaginal samples were colonized mostly by *Lactobacillus* (68.38%), *Prevotella* (7.37%), *Gardnerella* (5.69%), *Megasphaera* (5.23%), *Olsenella* (3.41%), *Sneathia* (2.69%) and others represented in **Fig. 5b**.

### Microbiota compositions from semen samples with leucocytes

We compared the microbiota compositions of 9 semen samples with leucocytes (2+ to 3++) and 9 semen samples without the presence of leucocytes. Semen samples with leucocytes tend to be significantly less colonized by *Lactobacillus reuteri* group, *Faecalibacterium* and more inhabited by *Bacteroides*, and *Prevotella* (Supplementary **Fig. S6)**. The corresponding microbiota at genera level from vaginal samples is presented in Supplementary **Fig. S7** showing *Lactobacillus* and *Gardnerella* as the most relative abundance. At the species taxonomic level, semen samples with leucocytes had more relative abundance of *Lactobacillus iners* and *Enterococcus faecium* compared with semen without leucocytes as shown in Supplementary **Fig. S8**. The corresponding female partners had *Lactobacillus iners* as the most abundant species as shown in Supplementary **Fig. S9**. Bacterial metabolic functional genes that were downregulated in the semen samples with leucocytes include but not limited to hemoglobin/transferrin/lactoferrin receptor protein, MFS transporter, OPA family, sugar phosphate sensor protein UhpC as presented in Supplementary **Table S1**.

### Microbiota compositions in couples with positive and negative IVF clinical outcome

This study compared the relative abundance of the microbiota in 12 couples with positive IVF clinical outcome and 24 couples with unsuccessful or negative IVF clinical outcome as shown in **Table 1a/1b**.

Semen samples with positive IVF clinical outcome have less alpha diversity as typified by Shannon index and phylogenetic diversity (Supplementary **Fig. S10**) and are significantly colonized by *Lactobacillus jensenii* group, *Faecalibacterium*, and significantly less colonized by *Proteobacteria* taxa, *Prevotella, Bacteroidetes* taxa, *Bacteroides* and lower *Firmicutes*/*Bacteroidetes* ratio compared with semen samples with negative IVF clinical outcome as shown in **Fig.6**.

The comparative proportion of the relative abundance at the species taxonomic level is represented in **Fig.7**.

LefSe comparison of the taxonomic microbiota biomarkers in the seminal fluid of men that had positive IVF clinical outcome and seminal fluid of those with negative IVF clinical outcome is presented in **Table 2**.

**Table 2:** LefSe comparison of the taxonomic biomarkers in the seminal fluid of men that had positive IVF clinical outcome and seminal fluid of those with negative IVF clinical outcome

| **Taxon name** | **LDA**<br>**effect size** | **p-value** | **p-value**<br>**(FDR)** | **Semen positive**<br>**IVF clinical**<br>**outcome** | **Semen negative IVF**<br>**clinical outcome** |
| --- | --- | --- | --- | --- | --- |
| *Cutibacterium* | 3.50568 | 0.00034 | 0.00034 | 0.76044 | 0.12494 |
| *Cutibacterium acnes* group | 3.45684 | 0.00067 | 0.00067 | 0.66262 | 0.09221 |
| *Propionibacteriales* | 3.48056 | 0.00068 | 0.00068 | 0.82203 | 0.23874 |
| *Propionibacteriaceae* | 3.49112 | 0.00068 | 0.00068 | 0.80718 | 0.21312 |
| *Salinifilum* | 3.70772 | 0.01579 | 0.01584 | 1.02006 | 0.00000 |
| *Leptotrichiaceae* | 3.12386 | 0.02997 | 0.03009 | 0.00000 | 0.26525 |
| *Facklamia hominis* group | 2.60266 | 0.02997 | 0.03011 | 0.00000 | 0.07949 |
| *Lactobacillus_uc* | 2.72540 | 0.03372 | 0.03390 | 0.01318 | 0.11892 |
| *Actinobacteria_c* | 4.76455 | 0.03899 | 0.03923 | 22.04688 | 10.41718 |
| *Micrococcales* | 2.99379 | 0.04751 | 0.04783 | 3.00235 | 2.80542 |
| *Streptomycetales* | 2.92289 | 0.04825 | 0.04861 | 0.16874 | 0.00170 |
| *Streptomycetaceae* | 2.92289 | 0.04825 | 0.04865 | 0.16874 | 0.00170 |
| *Streptomyces* | 2.92289 | 0.04825 | 0.04868 | 0.16874 | 0.00170 |
| *Cutibacterium avidum* | 2.44565 | 0.04825 | 0.04871 | 0.05682 | 0.00568 |
| *Saccharibacteria_TM7* | 2.20252 | 0.04825 | 0.04874 | 0.00000 | 0.03151 |
| *Saccharimonas_c* | 2.20252 | 0.04825 | 0.04878 | 0.00000 | 0.03151 |
| *Saccharimonas_o* | 2.20252 | 0.04825 | 0.04881 | 0.00000 | 0.03151 |
| *Saccharimonas_f* | 2.20252 | 0.04825 | 0.04885 | 0.00000 | 0.03151 |
| *Prevotella_uc* | 2.60734 | 0.04825 | 0.04888 | 0.00000 | 0.08053 |
| *Actinomyces europaeus*<br>group | 2.45905 | 0.04825 | 0.04891 | 0.00000 | 0.05622 |
| *Streptococcus_uc* | 2.02046 | 0.04825 | 0.04895 | 0.00000 | 0.01467 |

Note: **FDR**=False Discovery Rate; **LDA**= Linear Discriminant Analysis

In addition, the LefSe comparison of the taxonomic microbiota biomarkers for positive IVF clinical outcome for the couples are presented in Supplementary **Table S2**.

Vaginal samples with positive IVF clinical outcome are significantly colonized by *Lactobacillus gasseri* group, higher *Firmicutes/Bacteroidetes* ratio and significantly less colonized by *Bacteroides, Bacteroidetes* taxa, *Lactobacillus reuteri* group, *Lactobacillus iners*, and *Lactobacillus crispatus* as represented in **Fig.8**.

LefSe comparison of the taxonomic microbiota biomarkers in the vaginal samples of women that had positive IVF clinical outcome and vaginal samples of those with negative IVF clinical outcome is presented in **Table 3**.

**Table 3:** LefSe comparison of the taxonomic biomarkers in the vaginal swabs of women that had positive IVF clinical outcome and vaginal swabs of women with negative IVF clinical outcome.

| **Taxon name** | **LDA effect size** | ***p*-value** | ***p*-value (FDR)** | **Vagina positive IVF clinical outcome** | **Vagina negative IVF clinical outcome** |
| --- | --- | --- | --- | --- | --- |
| *Bacteroidetes* | 3.95843 | 0.00783 | 0.00783 | 0.01044 | 1.82702 |
| *Bacteroidia* | 3.95832 | 0.00889 | 0.00890 | 0.00978 | 1.82624 |
| *Bacteroidales* | 3.95832 | 0.00889 | 0.00891 | 0.00978 | 1.82624 |
| *Prevotellaceae* | 3.95382 | 0.01060 | 0.01064 | 0.00267 | 1.80030 |
| *Prevotella* | 3.95366 | 0.01060 | 0.01066 | 0.00267 | **1.79968** |
| *Citrobacter* | 2.17062 | 0.01579 | 0.01589 | 0.02942 | 0.00000 |
| *Citrobacter koseri* | 2.17062 | 0.01579 | 0.01591 | 0.02942 | 0.00000 |
| ***Cutibacterium acnes* group** | 3.04732 | 0.01822 | 0.01839 | 0.00000 | **0.22235** |
| *Enterobacterales_uc* | 2.34217 | 0.01864 | 0.01884 | 0.04385 | 0.00008 |
| *Prevotella timonensis* | 3.79064 | 0.01866 | 0.01888 | 0.00224 | **1.23701** |
| *Cutibacterium* | 3.06412 | 0.02560 | 0.02594 | 0.00038 | 0.23154 |
| *Streptococcus agalactiae* | 4.38488 | 0.03698 | 0.03767 | 8.66950 | 3.81779 |
| *Veillonella* | 2.28915 | 0.04785 | 0.04881 | 0.02708 | 0.06579 |
| ***Fusobacteria*** | 2.40327 | 0.04825 | 0.04928 | 0.00000 | 0.05037 |
| ***Fusobacteria_c*** | 2.40327 | 0.04825 | 0.04935 | 0.00000 | 0.05037 |
| ***Fusobacteriales*** | 2.40327 | 0.04825 | 0.04942 | 0.00000 | 0.05037 |
| ***Mobiluncus*** | 2.52442 | 0.04825 | 0.04962 | 0.00000 | 0.06573 |
| ***Mobiluncus curtisii* group** | 2.51681 | 0.04825 | 0.04968 | 0.00000 | 0.06468 |
| ***Veillonella dispar*** | 2.37510 | 0.04825 | 0.04975 | 0.00000 | 0.04684 |

Note: **FDR**=False Discovery Rate; **LDA**= Linear Discriminant Analysis

The results from PICRUSt indicated that some bacterial metabolic functional genes were upregulated in the semen of men who had positive IVF clinical outcome. For example, bacterial metabolic functional gene orthologs for phenylalanyl-tRNA synthetase beta chain was significantly upregulated (*P*=0.0231) with LDA effect size of 2.0847, when compared with bacterial metabolic genes from seminal fluid of men with negative IVF clinical outcome. Other several bacterial metabolic functional gene orthologs that were significantly upregulated include but not limited to methionyl aminopeptidase (P=0.0231), peptide/nickel transport system permease protein (P=0.0132), chaperonin GroEL (P**=**0.0286), glucose-6-phosphate 1-dehydrogenase (P=0.0374), mycothione reductase (P=0.0318), multicomponent Na+:H+ antiporter subunit D, E, G, A, F and subunit C were significantly upregulated (Supplementary **Table S3**).

Similarly, bacterial metabolic functional genes in the vagina of women with positive IVF clinical outcome were upregulated. Notably, iron/zinc/manganese/copper transport system permease protein (P=0.0326) and diphosphoinositol-polyphosphate diphosphatase (P=0.0369) had a 2-fold increase with positive IVF clinical outcome when compared with negative IVF clinical outcome (**Table 4**).

**Table 4:**
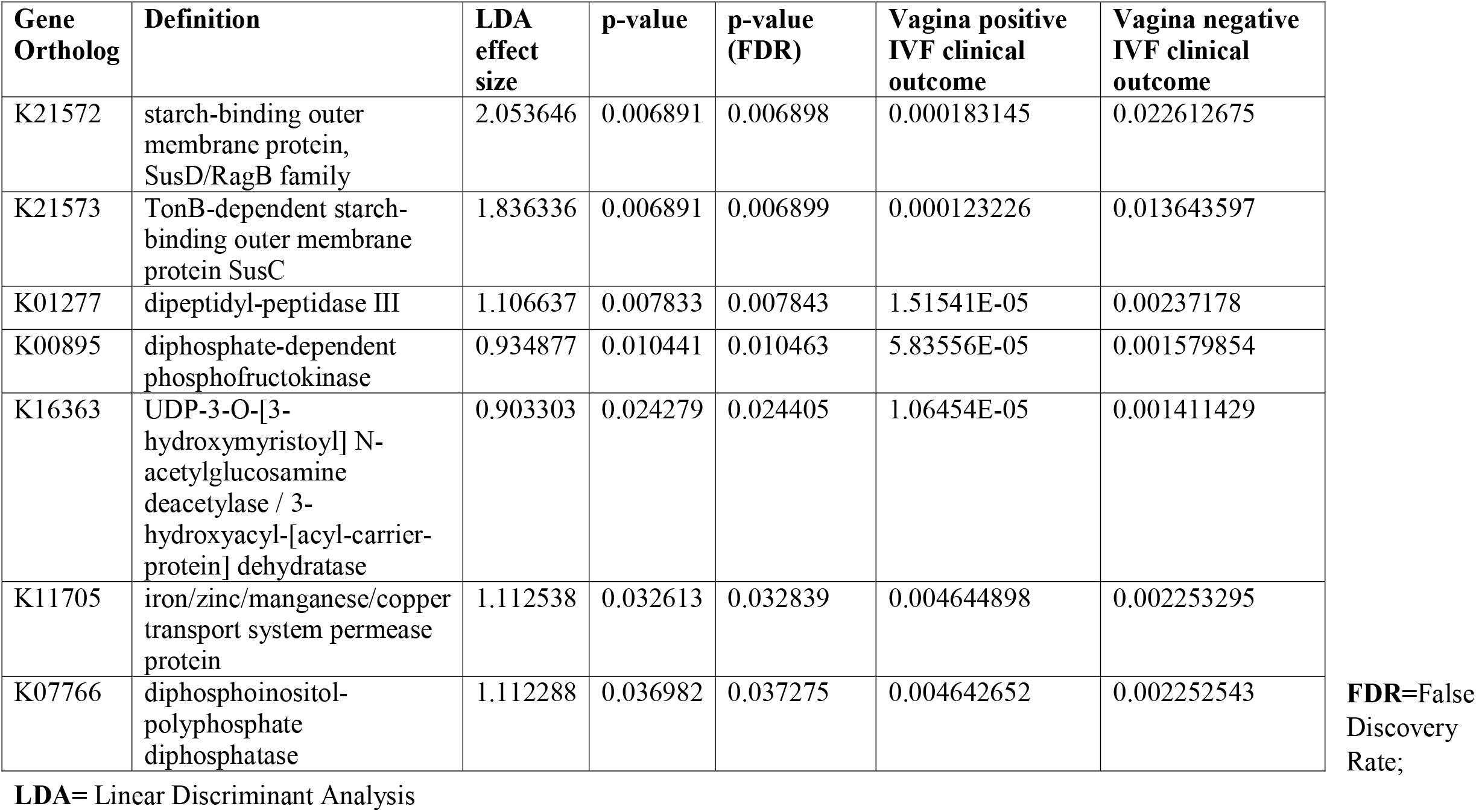
Bacterial metabolic functional gene orthologs in the vagina of women with positive IVF clinical outcome and women with negative IVF clinical outcome.

| **Gene Ortholog** | **Definition** | **LDA effect size** | **p-value** | **p-value (FDR)** | **Vagina positive IVF clinical outcome** | **Vagina negative IVF clinical outcome** |
| --- | --- | --- | --- | --- | --- | --- |
| K21572 | starch-binding outer membrane protein, SusD/RagB family | 2.053646 | 0.006891 | 0.006898 | 0.000183145 | 0.022612675 |
| K21573 | TonB-dependent starch-binding outer membrane protein SusC | 1.836336 | 0.006891 | 0.006899 | 0.000123226 | 0.013643597 |
| K01277 | dipeptidyl-peptidase III | 1.106637 | 0.007833 | 0.007843 | 1.51541E-05 | 0.00237178 |
| K00895 | diphosphate-dependent phosphofructokinase | 0.934877 | 0.010441 | 0.010463 | 5.83556E-05 | 0.001579854 |
| K16363 | UDP-3-O-[3-hydroxymyristoyl] N-acetylglucosamine deacetylase / 3-hydroxyacyl-[acyl-carrier-protein] dehydratase | 0.903303 | 0.024279 | 0.024405 | 1.06454E-05 | 0.001411429 |
| K11705 | iron/zinc/manganese/copper transport system permease protein | 1.112538 | 0.032613 | 0.032839 | 0.004644898 | 0.002253295 |
| K07766 | diphosphoinositol-polyphosphate diphosphatase | 1.112288 | 0.036982 | 0.037275 | 0.004642652 | 0.002252543 |

**FDR**=False Discovery Rate;
**LDA**= Linear Discriminant Analysis

## Discussion

From our knowledge, this is the first study from Nigeria that utilized next-generation sequencing (NGS) technology to determine the microbiota compositions of the seminal fluids and vaginal swabs from couples seeking assisted reproductive health care. The results, taken together revealed that the semen microbiota is highly polymicrobial in nature, typified by alpha diversity indexes such as Shannon index and phylogenetic diversity but low in species concentrations, as shown in similar study by Mandar et al.^**26**^. This finding was consistent with our previous pilot study of seminal fluids in a tertiary hospital that revealed varying bacterial diversities that are unique in each sample in contrast to culture-dependent methods^**17**^. The origin of these diverse bacteria in seminal fluids is still not fully determined, but interestingly most of the bacterial species are closely related to human vaginal microbes^**^18, 27, 28^**^, urine microbiota^**29**^ and urethra^**30**^. In contrast, the vaginal microbiota had higher species concentrations with less bacterial diversity. The family taxa, *Lactobacillaceae* and the *Lactobacillus* genus was significantly higher in the vagina compared with the semen (61.74% vs 43.86%) which is consistent with several studies showing that the healthy vagina is colonized by *Lactobacilli* that helps to fend off pathogenic microbes by increasing the pH, and preventing urogenital infections^**31**^. However, previous studies have shown that *Lactobacillus* is part of a normal microbiota of the seminal fluid in healthy subjects^**32**^. This study shows that *Gardnerella* (25.45% vs 5.86%) and *Veillonella* (7.78% vs 0.04%) were more in abundance in the seminal fluids with normospermia than the corresponding vagina microbiota, although *Gardnerella vaginalis* has severally been associated with bacterial vaginosis^**18**^. The physiological role of *Gardnerella vaginalis* in normospermic healthy subjects is yet to be determined as Weng et al^**33**^ found that *Lactobacillus, Gardnerella, Propionibacterium* and *Atopobium* were relatively more in abundance and significantly present in the normal semen samples. In this study, we found that relative signatures of bacterial communities could be used to disentangle semen categories. Men seeking reproductive health care in the tested population, though found to be normospermic tend to have more *Lactobacillus* >*Gardnerella* >*Veillonella* >*Corynebacterium*, while the female partners have more *Lactobacillus* >*Prevotella* >*Gardnerella* >*Streptococcus*. This suggest that the source of their infertility could probably be more than altered bacterial communities. A similar finding was reported by Hou *et al* ^**34**^ showing that infertile subjects did not have altered or unusual semen bacterial communities compared to normal sperm donors. The men categorized as oligospermic tend to have more *Prevotella* >*Escherichia* > *Lactobacillus* >*Shuttleworthia* >*Serratia* >*Megasphaera* >*Gardnerella* >*Sneathia* and their female partners tend to have more *Lactobacillus* >*Streptococcus* >*Gardnerella* >*Lactococcus* >*Bifidobacterium* >*Prevotella*. It appears that oligospermic men may be under the influence of these pathogens that overwhelms *Lactobacillus* protective activities. The factors responsible for these microbial community differences observed in oligospermic couples are yet to be delineated. In contrast, azoospermic men in these cohorts were observed to have more *Lactobacillus* >*Enterococcus* >*Corynebacterium* >*Veillonella* >*Gardnerella* >*Ureaplasma* >*Prevotella* while their female partners have more *Lactobacillus* >*Prevotella* >*Gardnerella* >*Megasphaera* >*Olsenella* >*Sneathia* >*Peptoniphillus* and other BV-associated bacteria. Previous studies have reported that several bacteria, including *Lactobacillus iners, Gardnerella vaginalis, Escherichia faecalis, E. coli* and *Staphylococcus aureus*, are associated with male infertility^**3**,**35**^. These bacterial taxa are common to both semen and vagina. The caveat is that they occur at different proportions in semen and vaginal niche. The differences in these bacterial communities in these partners may be due to sexual intercourse, episodes of receptive oral sex, anal sex before vaginal intercourse which have been reported to influence vaginal and genital tract microbiota in infertile couples^**36**^. The presence of leucocytospermia or pyospermia may have been triggered by the presence of *Gardnerella, Prevotella* and other BV-associated bacteria which leads to alteration in the partner’s vaginal microbiota, and BV has been associated with a 40% increase in the risk of Pre-term birth^**37**^, and infertility^**4**^. The occurrence of BV-related microbiota in semen suggests a possible reservoir and supports the concept of sexual transmission of BV, besides semen’s alkaline properties, which may alter the acidic pH of the vagina leading to BV^**38**^. An earlier study examined adherence, biofilm formation, and cytotoxicity *in vitro* for *G. vaginalis* strains isolated from women with BV as well as other BV-associated bacteria, including *Atopobium, Prevotella*, and *Mobiluncus*^**39**^. In terms of the impact of BV on IVF, several authors have observed high rates of BV on women with reproductive IVF failure and adverse pregnancy outcome^**40**,**41**,**42**^ which corroborates with the findings in this study. The negative IVF clinical outcome observed in these cohorts of men and women may have been due to induction of inflammatory response that inhibited the sperms from fertilizing the ovum. The microbiota biomarkers identified in women with negative IVF clinical outcome points towards an infectious perturbation of the IVF process. For example, in this study, the significant increase of *Bacteroidales, Prevotellaceae, Mobiluncus curtisii* and *Cutibacterium acnes* in the vagina of those with negative IVF clinical outcome lends credence to previous observations on causes of IVF failure^**4**,**5**^. *Cutibacterium acnes* are involved in the inflammation of the skin by secreting lipase enzymes which metabolize sebum into free fatty acids, but its activity in the vagina of women with negative IVF clinical outcome is yet to be determined. A recent study has demonstrated that IVF does not occur in a sterile environment and presence of *Staphylococcus sp*. and *Alphaproteobacteria* are associated with clinical indicators such as sperm and embryo quality^43^.

Transferrin/lactoferrin receptor proteins were significantly upregulated in the bacterial metabolic genes found in semen samples without leucocytes and with positive IVF clinical outcome when compared with those that had negative IVF clinical outcome. It should be noted that lactoferrin is a member of the iron-binding transferrin proteins known to have antimicrobial properties and plays a significant role in mucosal immune response. In addition, lactoferrin aid neutrophils by regulating hydroxyl radical production and participates in the secretion of IgA antibodies^**44**^.

## Conclusions

For the fact that semen samples of men with positive IVF clinical outcome were significantly colonized by *Lactobacillus jensenii* group, *Faecalibacterium*, and significantly less colonized by *Proteobacteria* taxa, *Prevotella, Bacteroidetes* taxa, suggest an association between semen and vaginal microbiota and a correlation to clinical IVF outcomes. In addition, this study has opened a window of possibility of using clinically tested probiotics therapy for men and women prior to IVF treatment. The need for implementing this approach has been advocated^45, 46^.

## Methods

### Ethical approval

The study was approved by the Ethics Review Committee on Human Research from Nnamdi Azikiwe University Teaching Hospital (**Ref # NAUTH/CS/66/VOL11/175/2018/111**). Participation in the study was voluntary. Informed written consent was obtained from the patients. All methods were performed in accordance with the **relevant guidelines and regulations**.

#### Selection criteria

Couples seeking reproductive health care at Nnamdi Azikiwe University Teaching Hospital (NAUTH), Nnewi campus, Anambra State, Nigeria, were referred to Life Fertility Center, Nnewi, for IVF/Embryo Transfer for self-cycle with cases of primary or secondary infertility after 1-12 years of uninterrupted sexual intercourse with partner.

#### Collection of specimens

Two high vaginal swabs were collected by a qualified Gynecologist with a non-lubricated sterile disposable plastic speculum and one of the swabs was agitated into a tube containing buffer for DNA preservation at ambient temperature and the other was used for microscopy to detect leucocytes. Each semen sample was produced by masturbation after 5 days of sexual intercourse abstinence and on the same day vaginal sample was collected.

#### Semen analysis

The semen quality of the patients was analyzed with Semen Quality Analyzer-Vision (SQA-V) Gold (Medical Electronic Systems, USA), following the manufacturer’s procedural instructions.

### Extraction of bacterial DNA from vaginal swabs/semen samples and Sequencing of the amplified 16S rRNA region

Bacterial DNA was extracted from the vaginal swabs/semen samples using a protocol developed by uBiome Inc. Briefly, samples were lysed using bead-beating, and DNA was extracted in a class 1000 clean room by a guanidine thiocyanate silica column-based purification method using a liquid-handling robot. PCR amplification of the 16S rRNA genes was performed with primers containing universal primers amplifying the V4 region (515F: GTGCCAGCMGCCGCGGTAA and 806R: GGACTACHVGGGTWTCTAAT). In addition, the primers contained Illumina tags and barcodes. Samples were barcoded with a unique combination of forward and reverse indexes allowing for simultaneous processing of multiple samples. PCR products were pooled, column-purified, and size-selected through microfluidic DNA fractionation. Consolidated libraries were quantified by quantitative real-time PCR using the Kapa Bio-Rad iCycler qPCR kit on a BioRad MyiQ before loading into the sequencer. Sequencing was performed in a pair-end modality on the Illumina NextSeq 500 platform rendering 2 x 150 bp pair-end sequences^**19**^.

#### 16S rRNA sequence analysis

Raw sequence reads were demultiplexed using Illumina’s BCL2FASTQ algorithm. Reads were filtered using an average Q-score > 30. The paired-end sequence FASTQ reads were imported into MG-RAST pipeline for quality check (QC). EzBiocloud Microbiome Taxonomic Profile (MTP) pipeline^**20**^ was employed for alpha and beta diversity estimation using PKSSU4.0 version database and Open reference UCLUST_MC2 for OTUs picking at 97% cut-off. Sequences were pre-screened using QIIME-UCLUST algorithms for at least 97% identity to ribosomal sequences from the RNA databases^**21**^. Rarefication to 1000 reads per sample was employed to calculate microbial diversity. Alpha-diversity was calculated for species richness by Abundance Coverage Estimate (ACE), Chao1 and Jackknife method, while diversity indexes were calculated by Shannon, Non-parametric Shannon and Simpson index. Principle coordinate analysis (PCoA) with Jensen-Shannon divergence distance metrices were used to evaluate beta diversity between vaginal and semen samples^**22**^. Linear discriminant analysis (LDA) effect size (LEfSe)^**23**^ was used to identify biologically and statistically significant differences in the OTU relative abundance. Phylogenetics Investigation of Communities by Reconstruction of Unobserved States (PICRUSt) was used to predict the metabolic function of the metagenomes from the 16S rRNA gene dataset^**24**^ with reference to Kyoto Encyclopedia of Genes and Genomes (KEGG) Orthologs categorizations^**25**^.

#### Availability of data and materials

The datasets used and or analyzed in the current study are available from the corresponding author on reasonable request.

## Data Availability

Availability of data and materials
The datasets used and or analyzed in the current study are available from the corresponding author on reasonable request.

## Acknowledgements

We would like to acknowledge uBiome Inc., San Francisco, California, USA, for assisting in the 16S rRNA metagenomics sequencing under an academic consortium grant-in-kind award to Dr. Kingsley Anukam. However, uBiome did not participate in the design of the study, collection, analysis, interpretation of data and in writing the manuscript. We sincerely thank all the couples who participated voluntarily in this study and lastly, we thank EZbiocloud™ for their bioinformatic pipeline used for analysis of some parts of this study.

## Author contributions

The author’s contributions were as follows; KCA designed the study and sourced for funding; SIO, NRA and KCA were responsible for taxonomic data organization, initial analysis and manuscript drafting; KCA was responsible for bioinformatic analysis; JII and SIO were involved in clinical evaluation and collection of samples; KCA, JII and NRA were the principal investigators and participated in the final design of the study, coordination and drafting the manuscript. All authors contributed to the interpretation of data and in writing and approval of the final manuscript.

## Additional information

### Competing interests

The authors declare no competing interests

### Ethics approval and consent to participate

Participation in the study was voluntary. Informed written consent was obtained from the patients. The study was approved by the Ethics Review Committee on Human Research from Nnamdi Azikiwe University Teaching Hospital (**Ref # NAUTH/CS/66/VOL11/175/2018/111**)

### Consent for publication

All authors agree to the submission of this article for publication.

## Supplementary Figure Legends

**Fig. S**1: Principle Co-Ordinate Analysis (PCoA) with the Bray-Curtis clustering in bacterial community diversities between semen and vagina

**Fig. S2:** The rarefaction curve showing the number of reads between semen samples and vaginal samples.

**Fig. S3:** Venn diagram showing number of taxa exclusive to normospermia, oligospermia, azoospermia, and taxa that were common to the semen categories.

**Fig. S4**. Comparative stacked relative (%) abundance of taxonomic phyla between semen and vagina in normospermia

**Fig. S5**. Comparative stacked relative (%) abundance of taxonomic genera between semen and vagina in normospermia.

**Fig. S6**. Comparative relative abundance of some selected taxa showing significant difference between semen samples without leucocytes and semen samples leucocytes

**Fig. S7:** Relative (%) abundance of taxonomic genera in the vaginal samples of the corresponding pyospermia samples

**Fig. S8**: Comparative relative (%) abundance of some species taxa between semen samples with leucocytes and semen without leucocytes.

**Fig. S9**: Relative (%) abundance of taxonomic species in the vaginal samples of the corresponding pyospermia samples

**Fig. S10:** Comparative alpha diversities between semen samples with positive IVF clinical outcome and semen samples with negative IVF clinical outcome.

## Notes

### Competing Interest Statement

The authors have declared no competing interest.

### Author Declarations

Ethical approval The study was approved by the Ethics Review Committee on Human Research from Nnamdi Azikiwe University Teaching Hospital (Ref # NAUTH/CS/66/VOL11/175/2018/111). Participation in the study was voluntary. Informed written consent was obtained from the patients. All methods were performed in accordance with the relevant guidelines and regulations.

