## Supplementary Figures FigS1-FigS10 for "Microbiota compositions from infertile couples seeking *in vitro* fertilization (IVF), using 16S rRNA gene sequencing methods: any correlation to clinical outcomes?"

**Fig.S1**

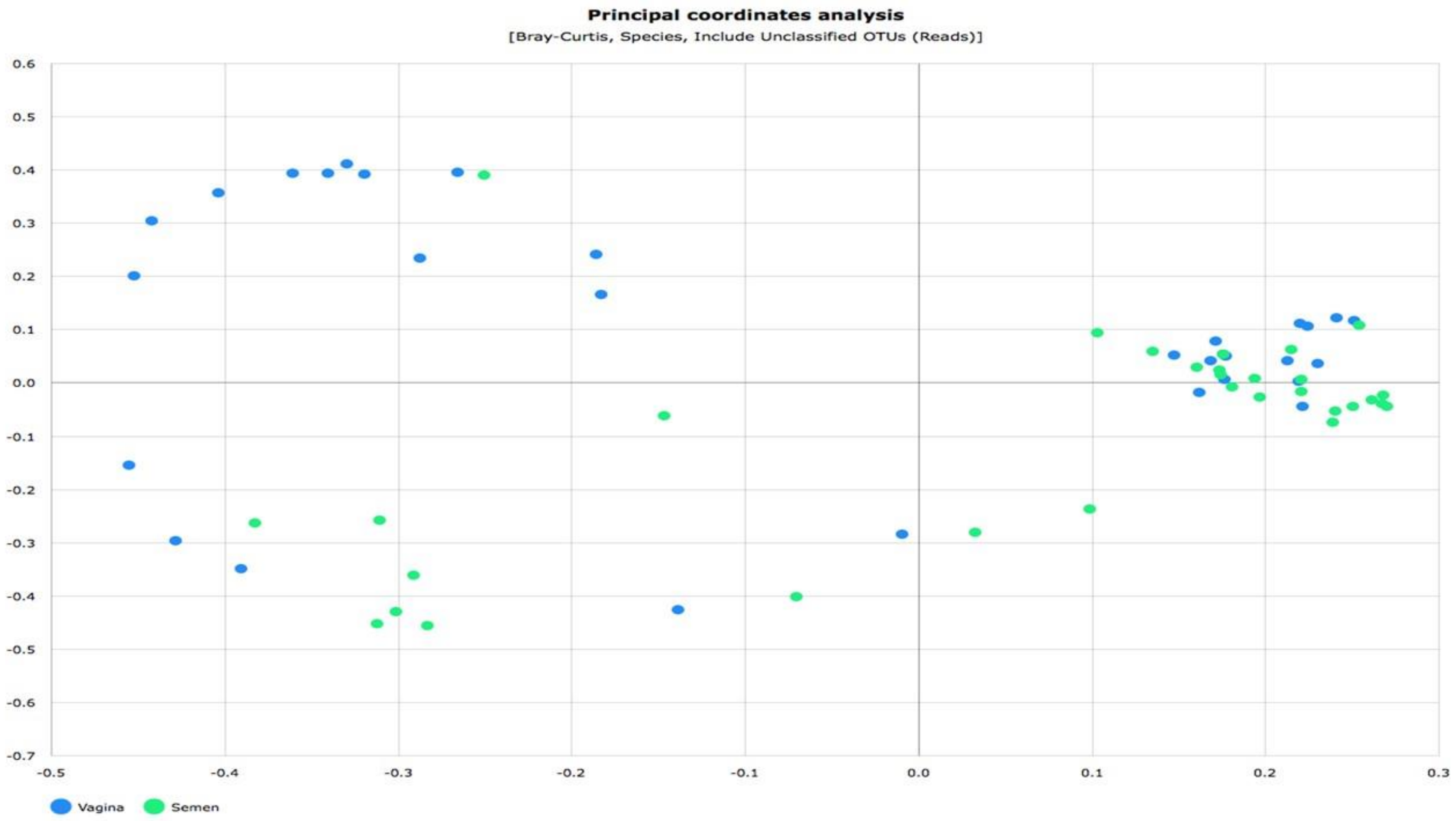

**Fig.S2**

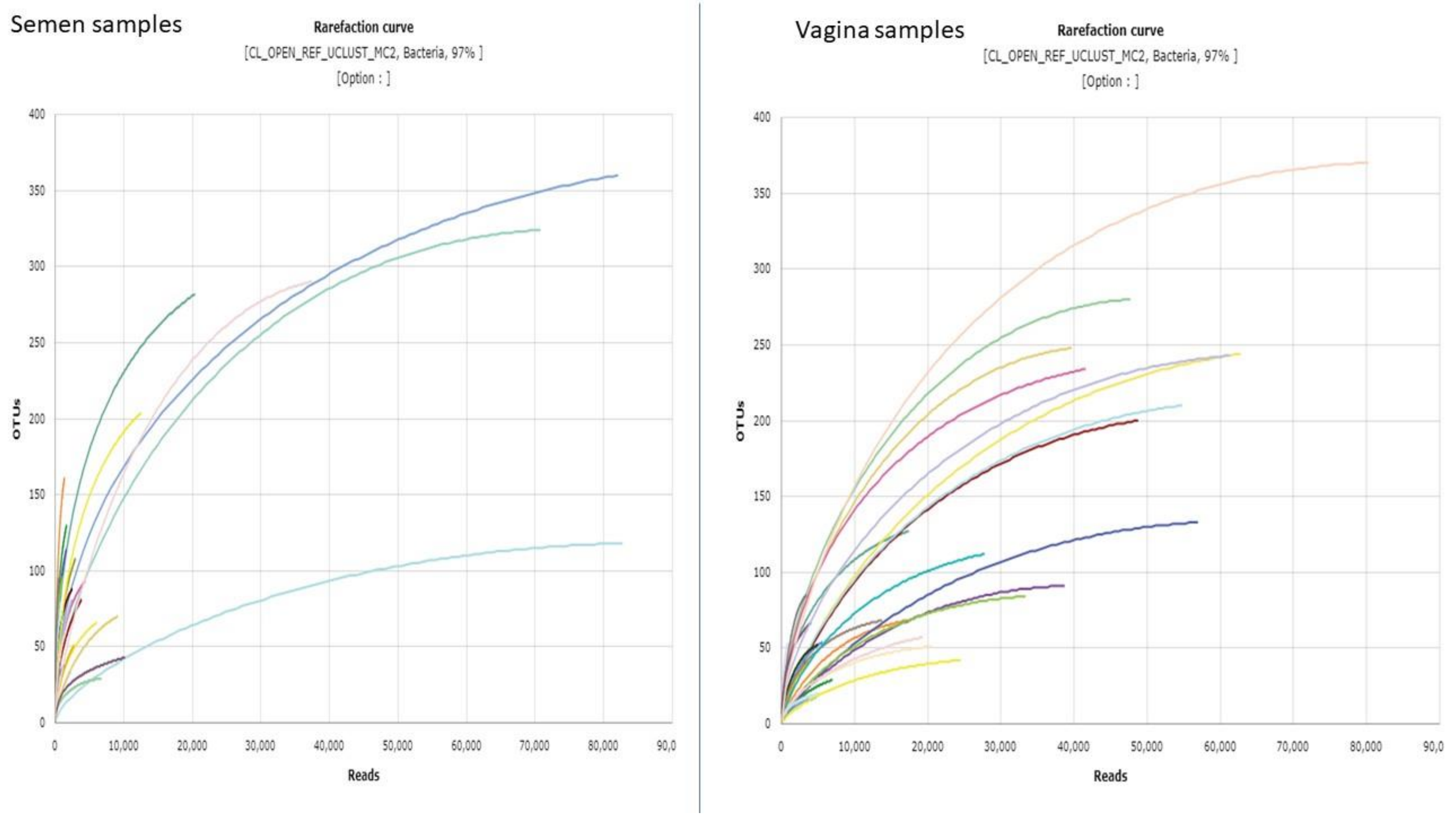
Semen samples
Rarefaction curve
[CL_OPEN_REF_UCLUST_MC2, Bacteria, 97% ]
[Option : ]
OTUs
400
350
300
250
200
150
100
50
0
0
10,000
20,000
30,000
40,000
50,000
60,000
70,000
80,000
90,0
Reads
Vagina samples
Rarefaction curve
[CL_OPEN_REF_UCLUST_MC2, Bacteria, 97% ]
[Option : ]
OTUs
400
350
300
250
200
150
100
50
0
0
10,000
20,000
30,000
40,000
50,000
60,000
70,000
80,000
90,0
Reads

Fig.S3

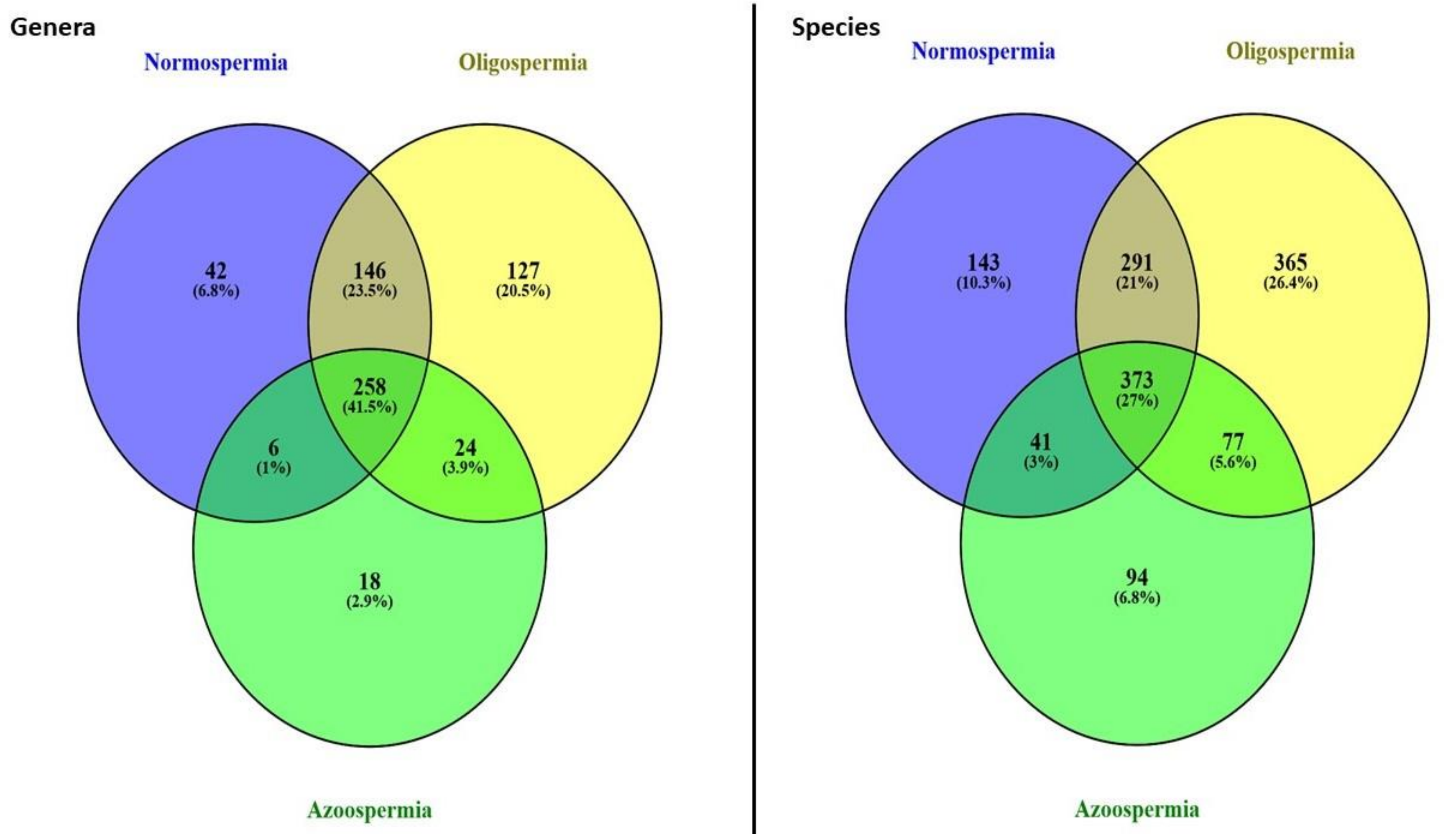

Genera
Normospermia
Oligospermia
42
(6.8%)
146
(23.5%)
127
(20.5%)
258
(41.5%)
6
(1%)
24
(3.9%)
18
(2.9%)
Azoospermia
Species
Normospermia
Oligospermia
143
(10.3%)
291
(21%)
365
(26.4%)
373
(27%)
41
(3%)
77
(5.6%)
94
(6.8%)
Azoospermia

**Fig.S4**

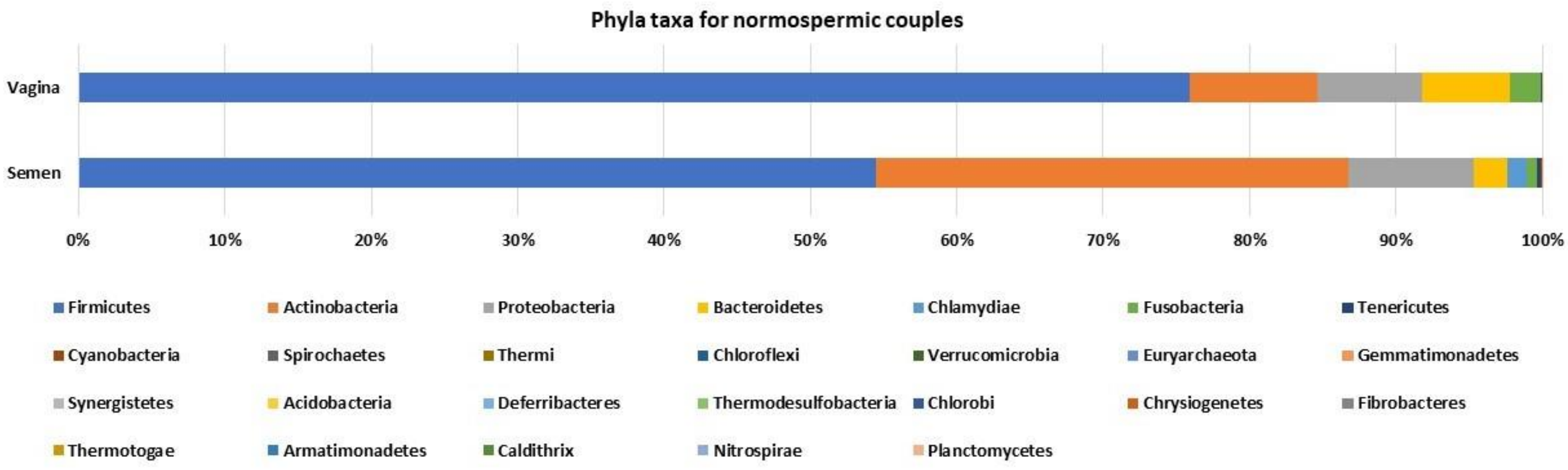

Phyla taxa for normospermic couples
Vagina
Semen
0%
10%
20%
30%
40%
50%
60%
70%
80%
90%
100%
Firmicutes
Actinobacteria
Proteobacteria
Bacteroidetes
Chlamydiae
Fusobacteria
Tenericutes
Cyanobacteria
Spirochaetes
Thermi
Chloroflexi
Verrucomicrobia
Euryarchaeota
Gemmatimonadetes
Synergistetes
Acidobacteria
Deferribacteres
Thermodesulfobacteria
Chlorobi
Chrysiogenetes
Fibrobacteres
Thermotogae
Armatimonadetes
Caldithrix
Nitrospirae
Planctomycetes

**Fig.S5**

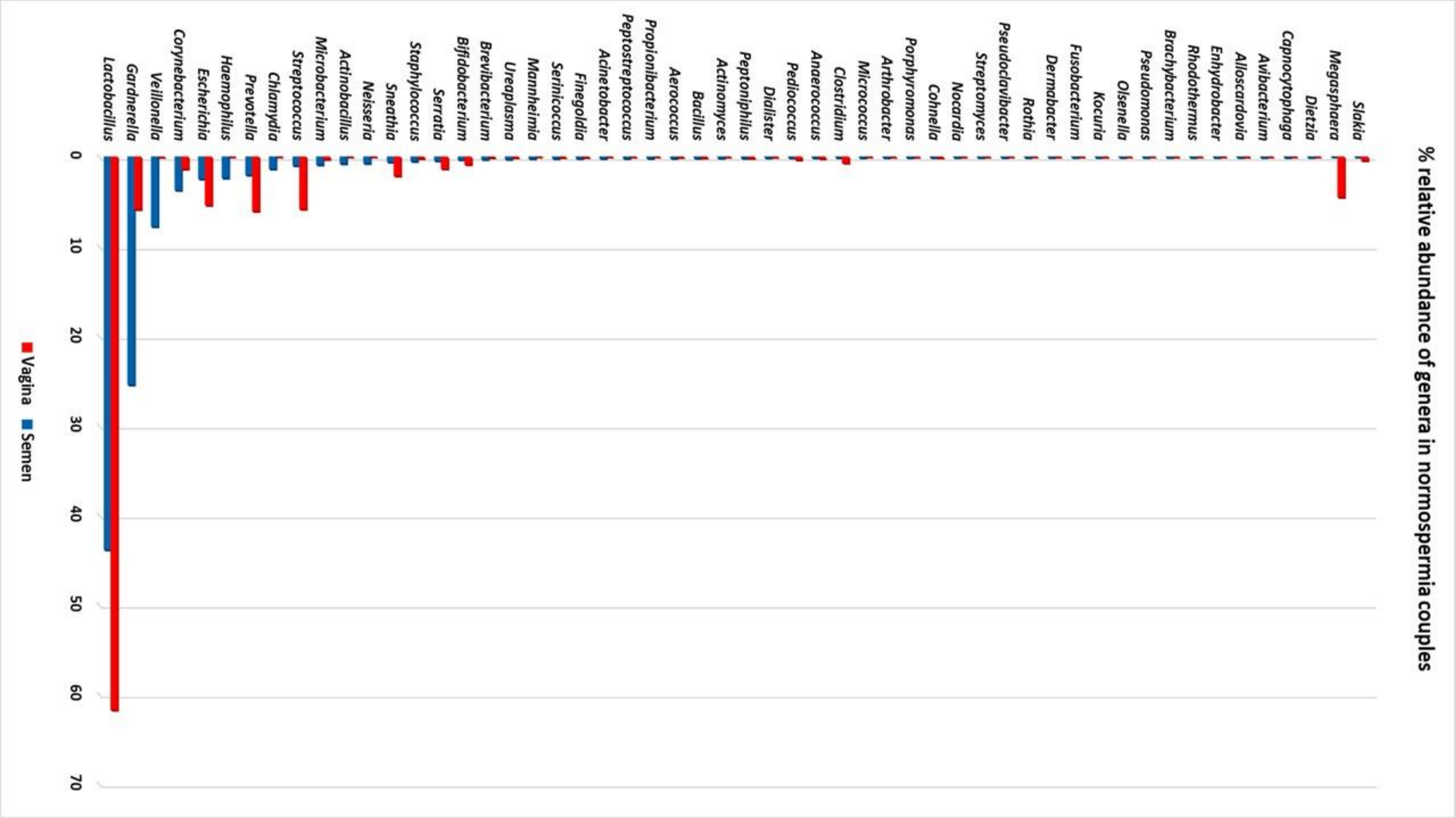
% relative abundance of genera in normospermia couples
Lactobacillus
Gardnerella
Veillonella
Corynebacterium
Escherichia
Haemophilus
Prevotella
Chlamydia
Streptococcus
Microbacterium
Actinobacillus
Neisseria
Sneathia
Staphylococcus
Serratia
Bifidobacterium
Brevibacterium
Ureaplasma
Mannheimia
Serinicoccus
Finegoldia
Acinetobacter
Peptostreptococcus
Propionibacterium
Aerococcus
Bacillus
Actinomyces
Peptoniphilus
Dialister
Pediococcus
Anaerococcus
Clostridium
Micrococcus
Arthrobacter
Porphyromonas
Cohnella
Nocardia
Streptomyces
Pseudoclavibacter
Rothia
Dermabacter
Fusobacterium
Kocuria
Olsenella
Pseudomonas
Brachybacterium
Rhodothermus
Enhydrobacter
Alloscardovia
Avibacterium
Capnocytophaga
Dietzia
Megasphaera
Slakia
0
10
20
30
40
50
60
70
Vagina
Semen

**Fig.S6**

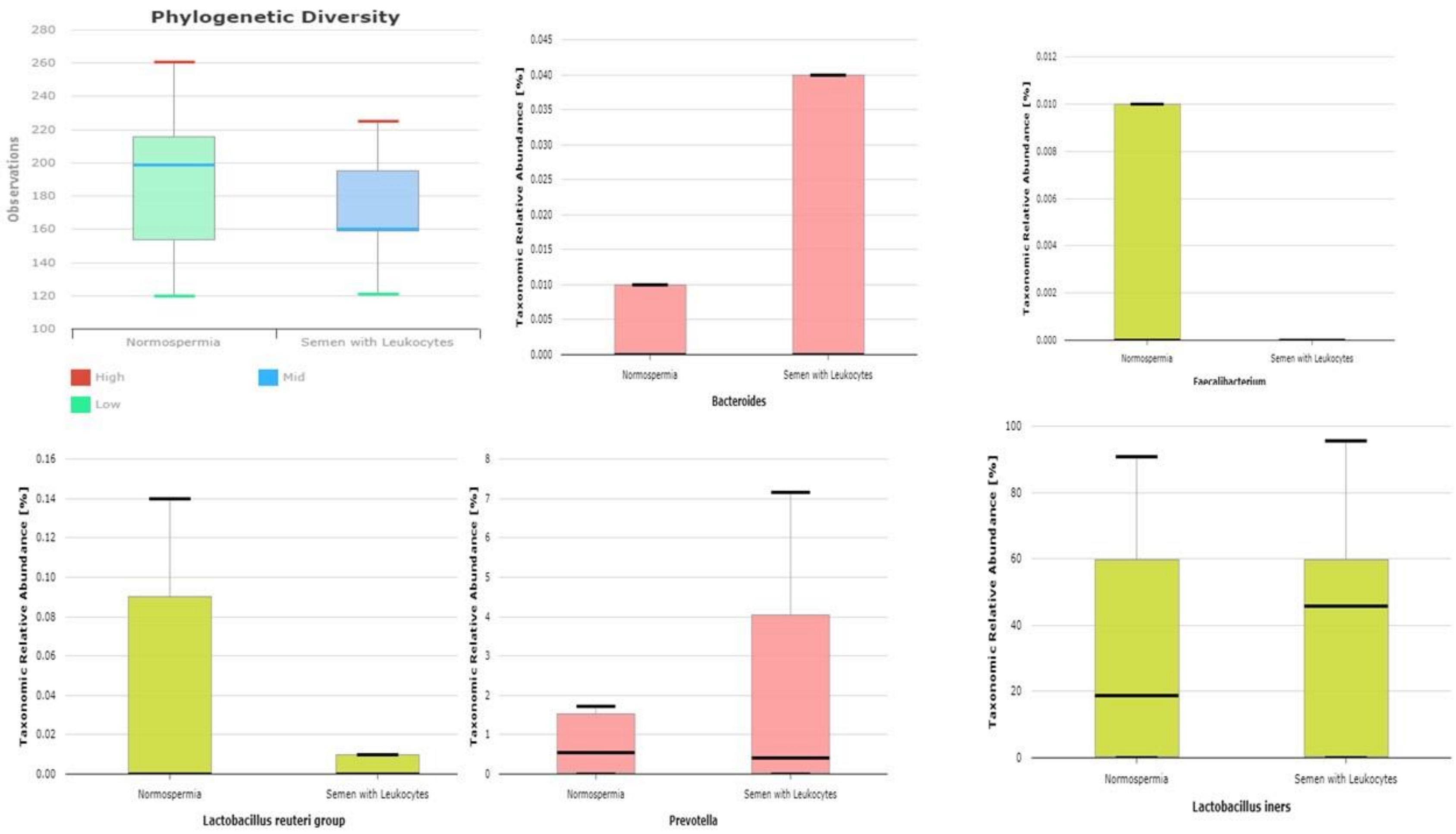

Phylogenetic Diversity
Observations
280
260
240
220
200
180
160
140
120
100
Normospermia
Semen with Leukocytes
High
Mid
Low
Taxonomic Relative Abundance [%]
Normospermia
Semen with Leukocytes
Bacteroides
Faecalibacterium
Lactobacillus reuteri group
Prevotella
Lactobacillus iners

**Fig.S7**

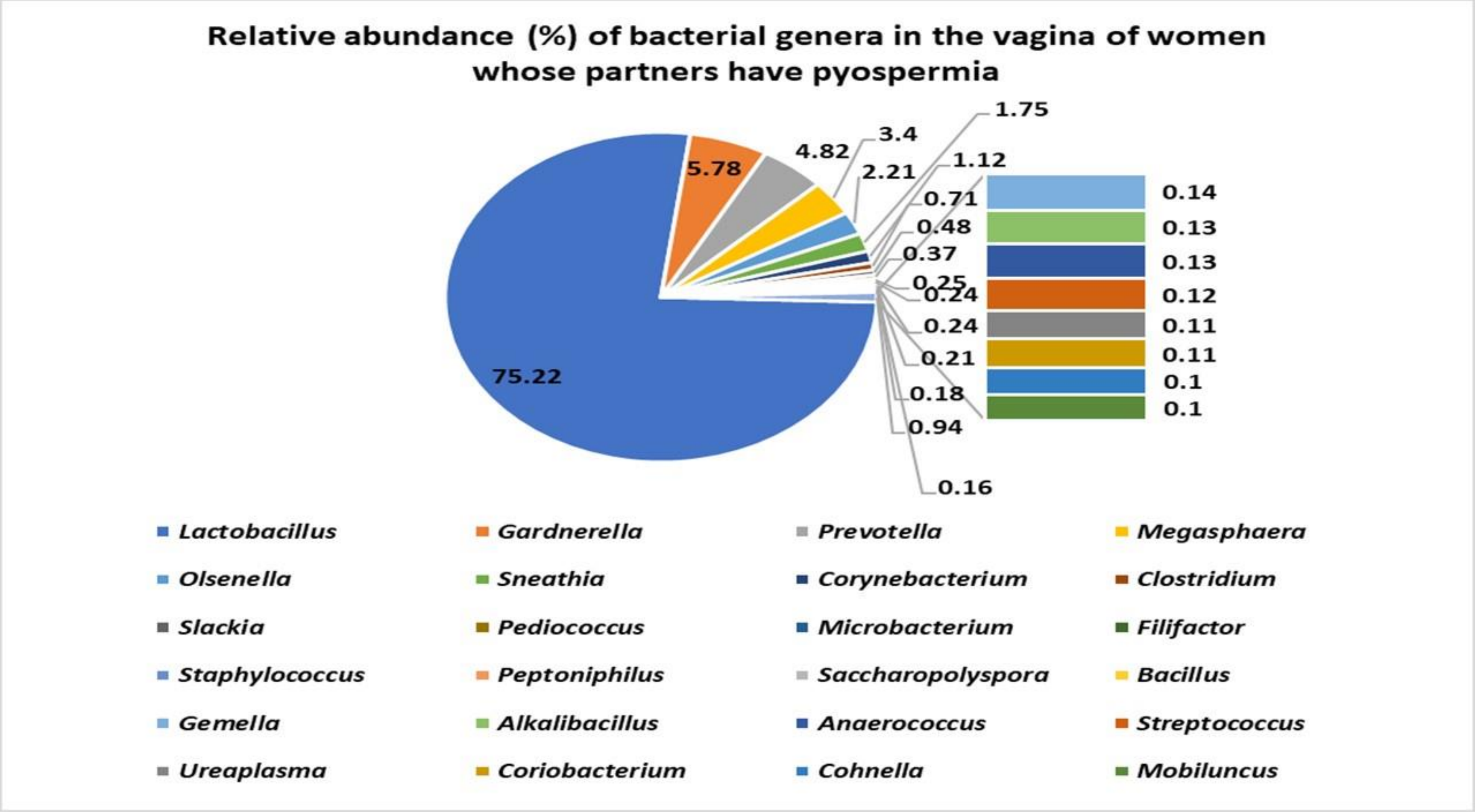
Relative abundance (%) of bacterial genera in the vagina of women whose partners have pyospermia
75.22
5.78
4.82
3.4
2.21
1.75
1.12
0.71
0.48
0.37
0.25
0.24
0.24
0.21
0.18
0.94
0.16
0.14
0.13
0.13
0.12
0.11
0.11
0.1
0.1
Lactobacillus
Gardnerella
Prevotella
Megasphaera
Olsenella
Sneathia
Corynebacterium
Clostridium
Slackia
Pediococcus
Microbacterium
Filifactor
Staphylococcus
Peptoniphilus
Saccharopolyspora
Bacillus
Gemella
Alkalibacillus
Anaerococcus
Streptococcus
Ureaplasma
Coriobacterium
Cohnella
Mobiluncus

**Fig.S8**

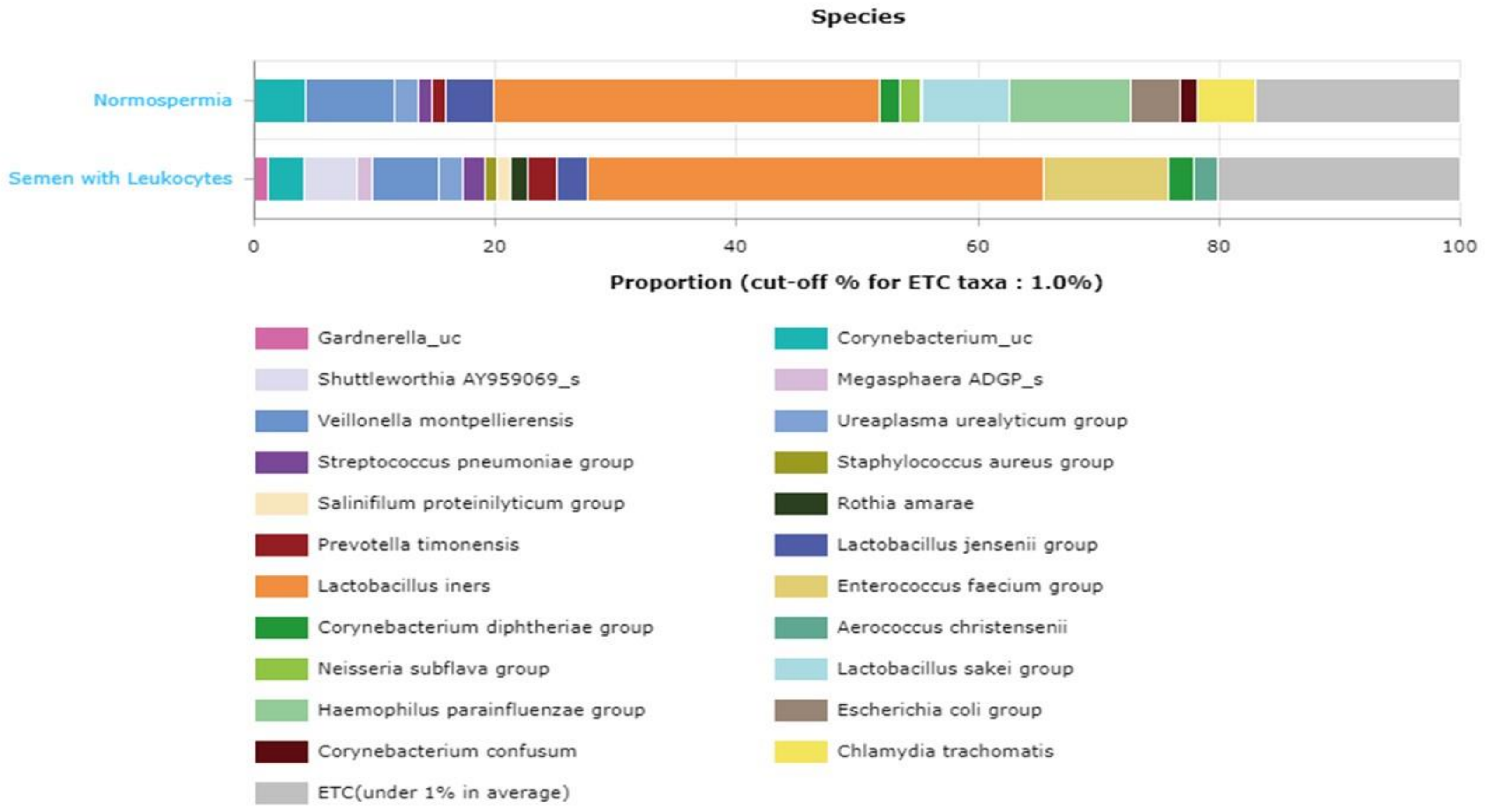
Species
Normospermia
Semen with Leukocytes
0
20
40
60
80
100
Proportion (cut-off % for ETC taxa : 1.0%)
Gardnerella_uc
Corynebacterium_uc
Shuttleworthia AY959069_s
Megasphaera ADGP_s
Veillonella montpellierensis
Ureaplasma urealyticum group
Streptococcus pneumoniae group
Staphylococcus aureus group
Salinifilum proteinilyticum group
Rothia amarae
Prevotella timonensis
Lactobacillus jensenii group
Lactobacillus iners
Enterococcus faecium group
Corynebacterium diphtheriae group
Aerococcus christensenii
Neisseria subflava group
Lactobacillus sakei group
Haemophilus parainfluenzae group
Escherichia coli group
Corynebacterium confusum
Chlamydia trachomatis
ETC(under 1% in average)

**Fig.S9**

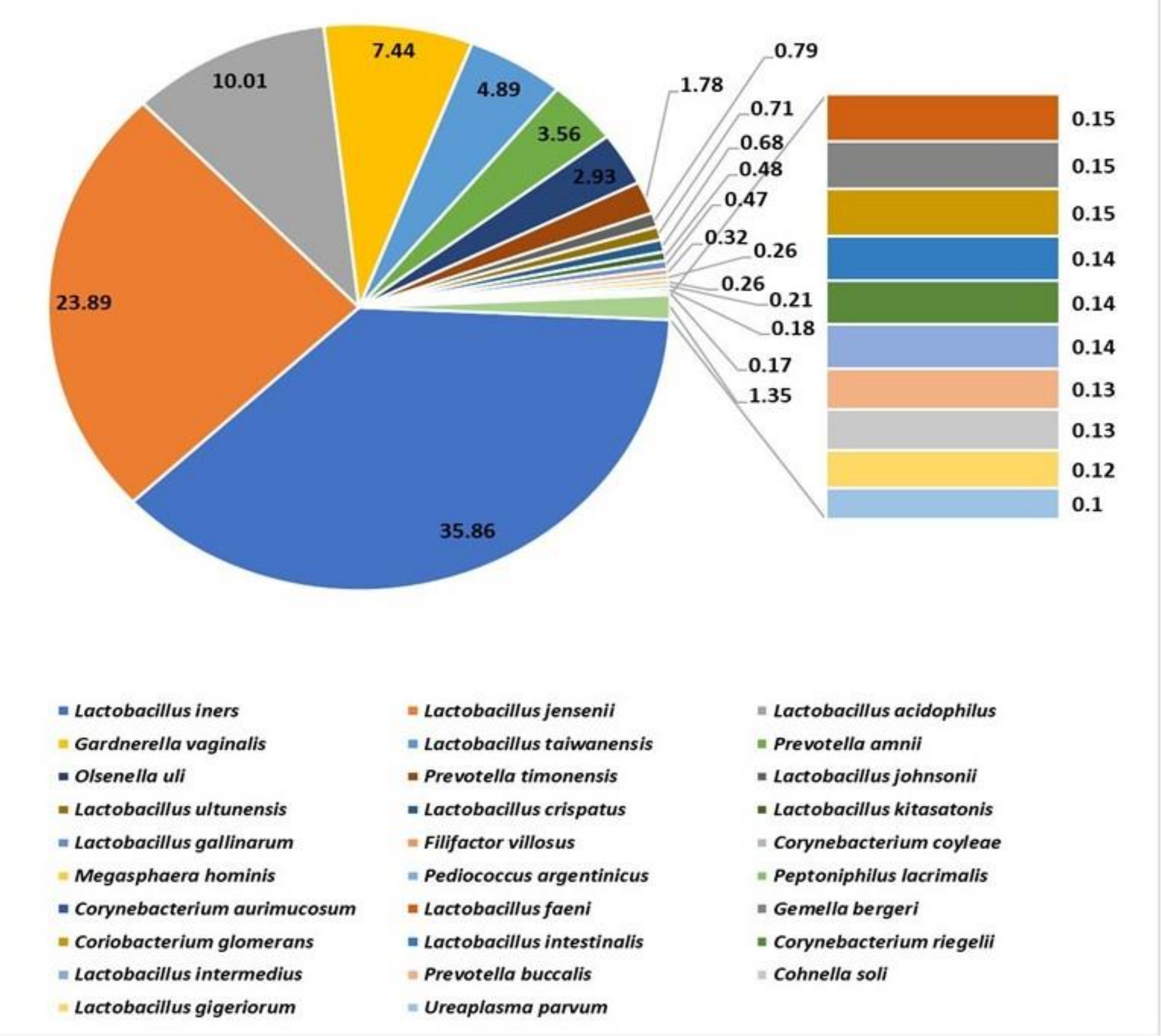
35.86
23.89
10.01
7.44
4.89
3.56
2.93
1.78
0.79
0.71
0.68
0.48
0.47
0.32
0.26
0.26
0.21
0.18
0.17
1.35
0.15
0.15
0.15
0.14
0.14
0.14
0.13
0.13
0.12
0.1
Lactobacillus iners
Lactobacillus jensenii
Lactobacillus acidophilus
Gardnerella vaginalis
Lactobacillus taiwanensis
Prevotella amnii
Olsenella uli
Prevotella timonensis
Lactobacillus johnsonii
Lactobacillus ultunensis
Lactobacillus crispatus
Lactobacillus kitasatonis
Lactobacillus gallinarum
Filifactor villosus
Corynebacterium coyleae
Megasphaera hominis
Pediococcus argentinicus
Peptoniphilus lacrimalis
Corynebacterium aurimucosum
Lactobacillus faeni
Gemella bergeri
Coriobacterium glomerans
Lactobacillus intestinalis
Corynebacterium riegelii
Lactobacillus intermedius
Prevotella buccalis
Cohnella soli
Lactobacillus gigeriorum
Ureaplasma parvum

**Fig.S10**

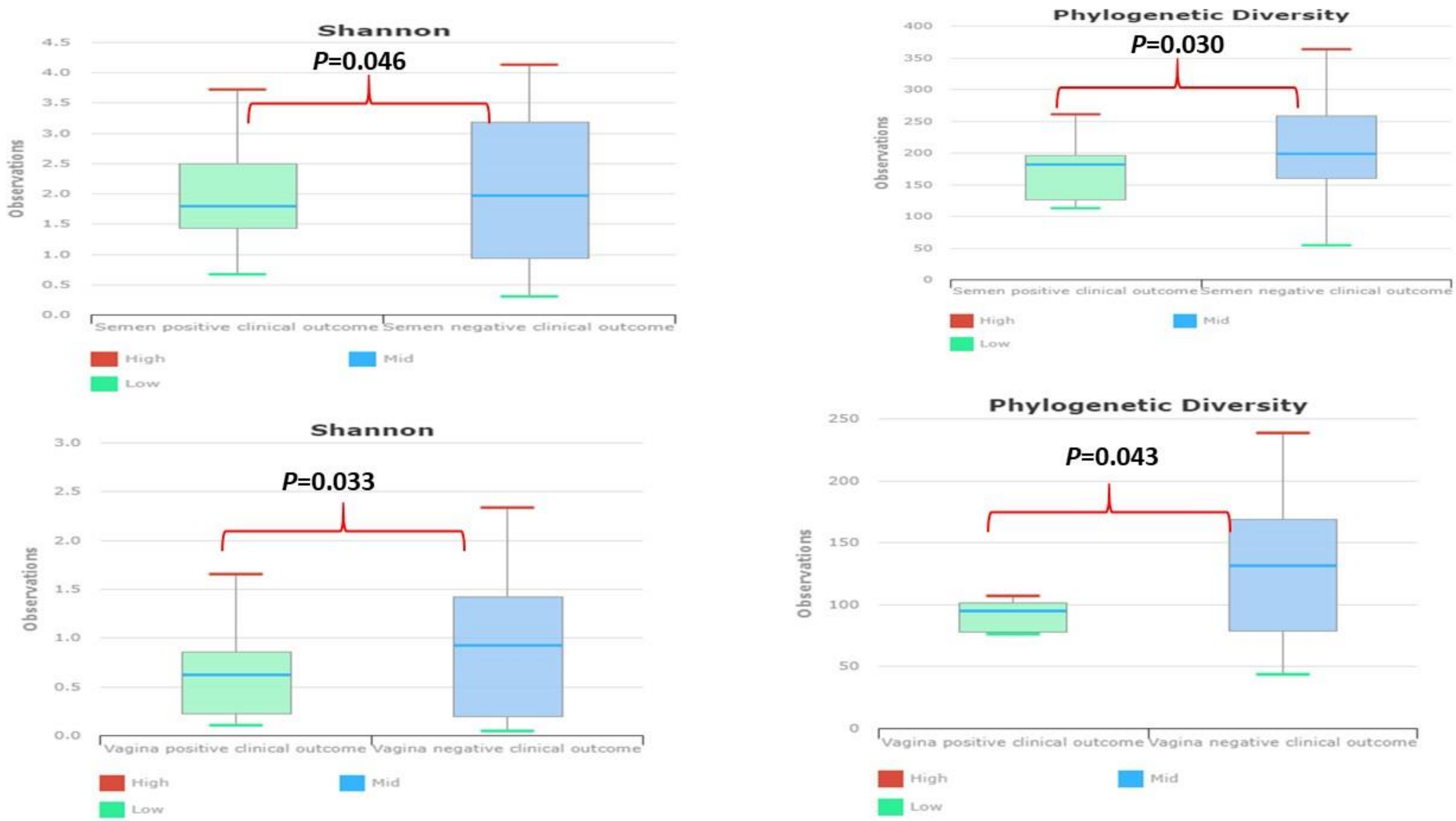
Shannon
P=0.046
Observations
4.5
4.0
3.5
3.0
2.5
2.0
1.5
1.0
0.5
0.0
Semen positive clinical outcome
Semen negative clinical outcome
High
Mid
Low
Phylogenetic Diversity
P=0.030
400
350
300
250
200
150
100
50
0
Shannon
P=0.033
3.0
2.5
2.0
1.5
1.0
0.5
0.0
Vagina positive clinical outcome
Vagina negative clinical outcome
Phylogenetic Diversity
P=0.043
250
200
150
100
50
0
