## Supplementary Tables 1-11 for "Microbiota compositions from infertile couples seeking *in vitro* fertilization (IVF), using 16S rRNA gene sequencing methods: any correlation to clinical outcomes?"

**Table S1:**

| Gene Ortholog | Metabolic Functional Gene Definition | LDA effect size | p-value | p-value (FDR) | Normospermia without leucocytes | Semen with Leukocytes |
|---|---|---|---|---|---|---|
| K02025 | multiple sugar transport system permease protein | 2.261725843 | 0.012724 | 0.012752175 | 0.064354389 | 0.100693286 |
| K00557 | tRNA (uracil-5-)-methyltransferase | 1.874490605 | 0.012724 | 0.012758059 | 0.015985129 | 0.001204857 |
| K16087 | hemoglobin/transferrin/lactoferrin receptor protein | 2.070152298 | 0.01991 | 0.020019188 | 0.025014425 | 0.001708239 |
| K02119 | V/A-type H+/Na+-transporting ATPase subunit C | 1.710055455 | 0.024682 | 0.024843508 | 0.002815807 | 0.012874332 |
| K10806 | acyl-CoA thioesterase YciA | 1.841834075 | 0.030413 | 0.030660244 | 0.017565963 | 0.003870828 |
| K07783 | MFS transporter, OPA family, sugar phosphate sensor protein UhpC | 1.722774099 | 0.030413 | 0.030702979 | 0.011095006 | 0.000731635 |
| K02122 | V/A-type H+/Na+-transporting ATPase subunit F | 1.697757071 | 0.030413 | 0.030712492 | 0.00294449 | 0.012716599 |
| K01995 | branched-chain amino acid transport system ATP-binding protein | 2.209931713 | 0.037253 | 0.037677881 | 0.09816811 | 0.130399195 |
| K03808 | paraquat-inducible protein A | 1.69974082 | 0.037253 | 0.037698353 | 0.010599677 | 0.000781914 |
| K03576 | LysR family transcriptional regulator, regulator for metE and metH | 1.87590023 | 0.037253 | 0.03770128 | 0.019489044 | 0.004660097 |
| K03604 | LacI family transcriptional regulator, purine nucleotide synthesis repressor | 1.949109778 | 0.045361 | 0.046168073 | 0.027013054 | 0.009424649 |
| K02806 | PTS system, nitrogen regulatory IIA component | 1.980305469 | 0.045361 | 0.046186102 | 0.026697121 | 0.007783834 |
| K03583 | exodeoxyribonuclease V gamma subunit | 1.886805413 | 0.045361 | 0.046240272 | 0.019449782 | 0.004238622 |
| K02453 | general secretion pathway protein D | 1.818376563 | 0.045361 | 0.046247505 | 0.017774012 | 0.004809466 |

**Table S2:**

| Taxon name | LDA effect size | p-value | p-value (FDR) | Semen positive IVF clinical outcome | Vagina positive IVF clinical outcome |
|---|---|---|---|---|---|
| Firmicutes | 5.09401 | 0.02782 | 0.03009 | 56.34075 | 81.17426 |
| Bacilli | 5.21938 | 0.01975 | 0.02116 | 47.3827 | 80.52727 |
| Lactobacillales | 5.33029 | 0.00949 | 0.00997 | 37.4993 | 80.28656 |
| Lactobacillaceae | 5.18136 | 0.04121 | 0.04617 | 32.84479 | 63.21107 |
| Lactobacillus | 5.18089 | 0.04121 | 0.04624 | 32.83665 | 63.16992 |
| Actinobacteria | 4.94624 | 0.00783 | 0.00817 | 22.27609 | 4.60474 |
| Actinobacteria_c | 4.96207 | 0.00428 | 0.0044 | 22.04688 | 3.71965 |
| Corynebacteriales | 4.84637 | 0.00643 | 0.00666 | 14.81763 | 0.7768 |
| Corynebacteriaceae | 4.84196 | 0.00643 | 0.00667 | 14.65991 | 0.76095 |
| Corynebacterium | 4.84186 | 0.00949 | 0.00999 | 14.64746 | 0.75181 |
| Bacillales | 4.68318 | 0.00783 | 0.00818 | 9.8834 | 0.2407 |
| Staphylococcaceae | 4.6722 | 0.01137 | 0.012 | 9.62297 | 0.22106 |
| Staphylococcus | 4.67058 | 0.00776 | 0.00809 | 9.57409 | 0.20708 |
| Staphylococcus aureus group | 4.61706 | 0.01137 | 0.01201 | 8.48793 | 0.20708 |
| Bacteroidetes | 4.58572 | 0.00134 | 0.00136 | 7.71487 | 0.01044 |
| Bacteroidia | 4.58477 | 0.0017 | 0.00173 | 7.69734 | 0.00978 |
| Bacteroidales | 4.58477 | 0.0017 | 0.00174 | 7.69734 | 0.00978 |
| Proteobacteria | 4.29491 | 0.00949 | 0.00996 | 6.98912 | 10.93298 |
| Corynebacterium_uc | 4.47953 | 0.01549 | 0.01653 | 6.16697 | 0.13377 |
| Porphyromonadaceae | 4.44654 | 0.04929 | 0.05568 | 5.59843 | 0.00656 |
| Porphyromonas | 4.44654 | 0.04929 | 0.05576 | 5.59843 | 0.00656 |
| Gammaproteobacteria | 4.43926 | 0.01369 | 0.01448 | 5.31457 | 10.81351 |
| Negativicutes | 4.38691 | 0.03556 | 0.03956 | 5.14508 | 0.27072 |
| Veillonellales | 4.38553 | 0.04183 | 0.04699 | 5.12968 | 0.27072 |
| Veillonellaceae | 4.38553 | 0.04183 | 0.04706 | 5.12968 | 0.27072 |

| | | | | | |
|---|---|---|---|---|---|
| Veillonella | 4.35812 | 0.00142 | 0.00144 | 4.58879 | 0.02708 |
| Pasteurellales | 4.21005 | 0.00685 | 0.00711 | 3.24644 | 0.00264 |
| Pasteurellaceae | 4.21005 | 0.00685 | 0.00712 | 3.24644 | 0.00264 |
| Haemophilus | 4.20881 | 0.00685 | 0.00713 | 3.23717 | 0.00264 |
| Haemophilus parainfluenzae group | 4.20555 | 0.00576 | 0.00596 | 3.21037 | 0 |
| Micrococcales | 4.15838 | 0.00039 | 0.00039 | 3.00235 | 0.12248 |
| Tissierellia | 4.04951 | 0.00181 | 0.00185 | 2.58313 | 0.34196 |
| Tissierellales | 4.04951 | 0.00181 | 0.00185 | 2.58313 | 0.34196 |
| Peptoniphilaceae | 4.04951 | 0.00181 | 0.00185 | 2.58313 | 0.34196 |
| Corynebacterium diphtheriae group | 4.03548 | 0.02629 | 0.02836 | 2.29907 | 0.12913 |
| Prevotellaceae | 4.00565 | 0.01103 | 0.01162 | 2.02863 | 0.00267 |
| Prevotella | 4.00166 | 0.03293 | 0.03659 | 2.01009 | 0.00267 |
| KV793764_s | 3.97497 | 0.03181 | 0.03497 | 1.8878 | 0 |
| Corynebacterium confusum | 3.94526 | 0.04901 | 0.05529 | 1.79477 | 0.03182 |
| Micrococcaceae | 3.77796 | 0.00124 | 0.00126 | 1.24546 | 0.0462 |
| Pseudomonadales | 3.72559 | 0.00086 | 0.00087 | 1.07523 | 0.0123 |
| Staphylococcus succinus group | 3.72754 | 0.01393 | 0.01481 | 1.06775 | 0 |
| Clostridia | 3.67147 | 0.03701 | 0.04123 | 0.97261 | 0.03431 |
| Clostridiales | 3.67147 | 0.03701 | 0.04129 | 0.97261 | 0.03431 |
| Betaproteobacteria | 3.61248 | 0.00027 | 0.00027 | 0.92971 | 0.11096 |
| Moraxellaceae | 3.64364 | 0.00086 | 0.00087 | 0.89077 | 0.01068 |
| Propionibacteriales | 3.61278 | 0.00005 | 0.00005 | 0.82203 | 0.00235 |
| Propionibacteriaceae | 3.60485 | 0.00005 | 0.00005 | 0.80718 | 0.00235 |
| Acinetobacter | 3.58208 | 0.00328 | 0.00336 | 0.77442 | 0.01068 |
| Cutibacterium | 3.57999 | 0.00004 | 0.00004 | 0.76044 | 0.00038 |
| Burkholderiales | 3.49195 | 0.00035 | 0.00035 | 0.72768 | 0.10832 |
| Finegoldia | 3.48308 | 0.00112 | 0.00113 | 0.71095 | 0.10429 |
| Brevibacteriaceae | 3.51418 | 0.02031 | 0.02184 | 0.69968 | 0.04665 |
| Brevibacterium | 3.51418 | 0.02031 | 0.02187 | 0.69968 | 0.04665 |
| Prevotella timonensis | 3.52223 | 0.04039 | 0.04519 | 0.66734 | 0.00224 |

| | | | | | |
|---|---|---|---|---|---|
| Cutibacterium acnes group | 3.52039 | 0.00002 | 0.00002 | 0.66262 | 0 |
| Finegoldia magna | 3.4283 | 0.00088 | 0.00089 | 0.63434 | 0.10011 |
| Comamonadaceae | 3.42454 | 0.00344 | 0.00353 | 0.5877 | 0.05745 |
| Rothia | 3.46403 | 0.03293 | 0.03654 | 0.58267 | 0.00076 |
| Dermabacteraceae | 3.38255 | 0.00516 | 0.00531 | 0.49778 | 0.0156 |
| Veillonella ratti group | 3.34061 | 0.03181 | 0.03492 | 0.4378 | 0 |
| Delftia | 3.25927 | 0.0194 | 0.02073 | 0.39414 | 0.03163 |
| Delftia tsuruhatensis group | 3.25927 | 0.0194 | 0.02076 | 0.39414 | 0.03163 |
| Microbacteriaceae | 3.2494 | 0.00588 | 0.00608 | 0.36765 | 0.01404 |
| Micrococcus luteus group | 3.23728 | 0.04039 | 0.04513 | 0.34509 | 0.00016 |
| Acinetobacter haemolyticus group | 3.05244 | 0.03181 | 0.03464 | 0.22508 | 0 |
| Actinomycetales | 2.98079 | 0.00878 | 0.00919 | 0.19705 | 0.01071 |
| Actinomycetaceae | 2.98079 | 0.00878 | 0.00921 | 0.19705 | 0.01071 |
| Zimmermannella | 2.95452 | 0.03181 | 0.03459 | 0.17955 | 0 |
| Zimmermannella bifida | 2.95452 | 0.03181 | 0.03487 | 0.17955 | 0 |
| Veillonella dispar | 2.93584 | 0.01393 | 0.01483 | 0.17069 | 0 |
| Streptomycetales | 2.92707 | 0.03181 | 0.03445 | 0.16874 | 0 |
| Streptomycetaceae | 2.92707 | 0.03181 | 0.0345 | 0.16874 | 0 |
| Streptomyces | 2.92707 | 0.03181 | 0.03454 | 0.16874 | 0 |
| Yersiniaceae | 2.72263 | 0.02734 | 0.02953 | 0.1372 | 0.03354 |
| Intrasporangiaceae | 2.8171 | 0.01393 | 0.01475 | 0.12868 | 0 |
| Brachybacterium | 2.81245 | 0.00543 | 0.00559 | 0.12776 | 0.00153 |
| Brachybacterium faecium group | 2.81385 | 0.00543 | 0.0056 | 0.12776 | 0.00131 |
| Actinomyces | 2.8047 | 0.04552 | 0.05129 | 0.12566 | 0.00961 |
| Enhydrobacter | 2.76726 | 0.01393 | 0.01477 | 0.11636 | 0 |
| Enhydrobacter aerosaccus group | 2.76726 | 0.01393 | 0.01479 | 0.11636 | 0 |
| Yersinia | 2.7521 | 0.01978 | 0.02122 | 0.11307 | 0.00568 |
| Yersinia pestis group | 2.7521 | 0.01978 | 0.02124 | 0.11307 | 0.00568 |
| Pseudomonas stutzeri group | 2.63966 | 0.03181 | 0.03478 | 0.08686 | 0 |
| Mobiluncus | 2.55388 | 0.03181 | 0.03506 | 0.07139 | 0 |

| | | | | | |
|---|---|---|---|---|---|
| Mobiluncus curtisii group | 2.55388 | 0.03181 | 0.03511 | 0.07139 | 0 |
| Acinetobacter junii group | 2.56823 | 0.03181 | 0.03501 | 0.06841 | 0 |
| Brevibacterium iodinum group | 2.43642 | 0.03181 | 0.03468 | 0.05228 | 0 |
| Cutibacterium granulosum | 2.32763 | 0.03181 | 0.03473 | 0.041 | 0 |
| Ralstonia syzygii group | 2.56562 | 0.03181 | 0.03482 | 0.01404 | 0 |
| Lactobacillus_uc | 3.46664 | 0.00174 | 0.00178 | 0.01318 | 0.59829 |
| Streptococcus agalactiae | 4.63698 | 0.01393 | 0.01485 | 0 | 8.6695 |
| Anaerococcus vaginalis group | 2.12563 | 0.03181 | 0.0352 | 0 | 0.02547 |

**Table S3:**

| Gene Ortholog | Definition | LDA effect size | p-value | p-value (FDR) | Semen positive IVF clinical outcome | Semen negative IVF clinical outcome |
|---|---|---|---|---|---|---|
| K01972 | DNA ligase (NAD+) | 2.127514 | 0.025784 | 0.027006 | 0.14999749 | 0.123372278 |
| K03111 | single-strand DNA-binding protein | 1.989716 | 0.047509 | 0.051646 | 0.11392864 | 0.094596717 |
| K02342 | DNA polymerase III subunit epsilon | 2.237197 | 0.020776 | 0.021533 | 0.113473107 | 0.079140673 |
| K03466 | DNA segregation ATPase FtsK/SpoIIIE, S-DNA-T family | 2.047748 | 0.013243 | 0.013504 | 0.107140416 | 0.085016167 |
| K01890 | phenylalanyl-tRNA synthetase beta chain | 2.084727 | 0.023163 | 0.02414 | 0.100749475 | 0.076641049 |
| K02335 | DNA polymerase I | 2.108086 | 0.002953 | 0.002964 | 0.10072175 | 0.075270088 |
| K01265 | methionyl aminopeptidase | 1.987601 | 0.023163 | 0.024148 | 0.10020196 | 0.080964916 |
| K01462 | peptide deformylase | 1.990179 | 0.043071 | 0.046367 | 0.09983316 | 0.080480433 |
| K07042 | probable rRNA maturation factor | 2.015016 | 0.038989 | 0.041653 | 0.099490781 | 0.078987216 |
| K02033 | peptide/nickel transport system permease protein | 2.129036 | 0.013243 | 0.013497 | 0.098642617 | 0.071923204 |
| K00655 | 1-acyl-sn-glycerol-3-phosphate acyltransferase | 2.028256 | 0.038989 | 0.041735 | 0.095686398 | 0.074541928 |
| K03070 | preprotein translocase subunit SecA | 1.942646 | 0.020776 | 0.021523 | 0.094463108 | 0.077137421 |
| K03177 | tRNA pseudouridine55 synthase | 2.079707 | 0.043071 | 0.046331 | 0.092473783 | 0.068644748 |
| K04077 | chaperonin GroEL | 2.139938 | 0.028658 | 0.030169 | 0.089787416 | 0.062383681 |
| K07058 | membrane protein | 2.047924 | 0.014856 | 0.015206 | 0.082515965 | 0.060382606 |
| K03665 | GTPase | 1.899305 | 0.03524 | 0.037399 | 0.082092403 | 0.066431247 |
| K01772 | protoporphyrin/coproporphyrin ferrochelatase | 2.052958 | 0.020776 | 0.021517 | 0.074863913 | 0.052470186 |
| K03327 | multidrug resistance protein, MATE family | 2.203422 | 0.047509 | 0.051642 | 0.073717078 | 0.105465668 |
| K01239 | purine nucleosidase | 2.108919 | 0.043071 | 0.046305 | 0.073311234 | 0.047810338 |

| | | | | | | |
|---|---|---|---|---|---|---|
| K02483 | two-component system, OmpR family, response regulator | 2.047107 | 0.03524 | 0.037454 | 0.07263009 | 0.050538717 |
| K00626 | acetyl-CoA C-acetyltransferase | 1.99962 | 0.03524 | 0.037419 | 0.069818706 | 0.05003624 |
| K00036 | glucose-6-phosphate 1-dehydrogenase | 1.975893 | 0.03524 | 0.037469 | 0.066485462 | 0.047765376 |
| K09167 | uncharacterized protein | 2.046138 | 0.038989 | 0.041791 | 0.045411383 | 0.023369705 |
| K02021 | putative ABC transport system ATP-binding protein | 1.958238 | 0.023163 | 0.024076 | 0.041531088 | 0.023564754 |
| K06919 | putative DNA primase/helicase | 2.013319 | 0.028658 | 0.03016 | 0.037171357 | 0.057594173 |
| K00656 | formate C-acetyltransferase | 2.070174 | 0.014856 | 0.015198 | 0.032817127 | 0.056124477 |
| K02022 | HlyD family secretion protein | 1.827474 | 0.025784 | 0.026946 | 0.032391741 | 0.019148527 |
| K00847 | fructokinase | 1.932501 | 0.011787 | 0.011997 | 0.031899598 | 0.048820644 |
| K07665 | two-component system, OmpR family, copper resistance phosphate regulon response regulator CusR | 1.979416 | 0.025784 | 0.026969 | 0.030221569 | 0.049095685 |
| K07636 | two-component system, OmpR family, phosphate regulon sensor histidine kinase PhoR | 1.949764 | 0.047509 | 0.051486 | 0.030145419 | 0.047760752 |
| K18851 | diacylglycerol O-acyltransferase / trehalose O-mycolyltransferase | 1.973155 | 0.049845 | 0.054394 | 0.028820919 | 0.010219762 |
| K00128 | aldehyde dehydrogenase (NAD+) | 1.915927 | 0.043071 | 0.046312 | 0.028683223 | 0.012403259 |
| K04069 | pyruvate formate lyase activating enzyme | 2.149586 | 0.006432 | 0.006486 | 0.027926333 | 0.055950172 |
| K05568 | multicomponent Na+:H+ antiporter subunit D | 1.927471 | 0.043071 | 0.04632 | 0.024558915 | 0.007835 |
| K05569 | multicomponent Na+:H+ antiporter subunit E | 1.921343 | 0.038989 | 0.04163 | 0.024144598 | 0.00765781 |
| K05571 | multicomponent Na+:H+ antiporter subunit G | 1.910777 | 0.038989 | 0.041709 | 0.023648463 | 0.007562758 |
| K05565 | multicomponent Na+:H+ antiporter subunit A | 1.916768 | 0.043071 | 0.046422 | 0.023639464 | 0.007327532 |
| K05570 | multicomponent Na+:H+ antiporter subunit F | 1.901026 | 0.038989 | 0.041657 | 0.023096116 | 0.007371991 |

| | | | | | | |
|---|---|---|---|---|---|---|
| K00516 | lytic starch monooxygenase | 1.907872 | 0.016639 | 0.017078 | 0.022382885 | 0.006405736 |
| K07741 | anti-repressor protein | 1.996787 | 0.016639 | 0.017098 | 0.021925715 | 0.04157828 |
| K05567 | multicomponent Na+:H+ antiporter subunit C | 1.852899 | 0.03524 | 0.037498 | 0.021176543 | 0.007122792 |
| K17883 | mycothione reductase | 1.837484 | 0.031803 | 0.033602 | 0.020921679 | 0.007364984 |
| K07007 | 3-dehydro-bile acid Delta4,6-reductase | 1.884045 | 0.031803 | 0.033694 | 0.020019634 | 0.035133099 |
| K06871 | uncharacterized protein | 2.007904 | 0.020776 | 0.021548 | 0.019971188 | 0.040138492 |
| K14059 | integrase | 2.082284 | 0.043071 | 0.046316 | 0.019184477 | 0.043156532 |
| K07175 | PhoH-like ATPase | 1.854199 | 0.016639 | 0.017104 | 0.019145187 | 0.00504872 |
| K04758 | ferrous iron transport protein A | 1.861288 | 0.028658 | 0.03012 | 0.01836619 | 0.032697865 |
| K03737 | pyruvate-ferredoxin/flavodoxin oxidoreductase | 1.871032 | 0.023163 | 0.024091 | 0.018221391 | 0.032882862 |
| K11904 | type VI secretion system secreted protein VgrG | 2.217267 | 0.013243 | 0.013514 | 0.017533436 | 0.050316935 |
| K07089 | uncharacterized protein | 1.82063 | 0.020776 | 0.021518 | 0.013720232 | 0.026753276 |
| K19337 | RpiR family transcriptional regulator, carbohydrate utilization regulator | 2.090217 | 0.028658 | 0.030141 | 0.013489406 | 0.037907035 |
| K12678 | autotransporter family porin | 1.967311 | 0.014856 | 0.015204 | 0.013421248 | 0.031771107 |
| K07484 | transposase | 2.53569 | 0.031803 | 0.033681 | 0.010953154 | 0.079415756 |
| K09812 | cell division transport system ATP-binding protein | 1.848876 | 0.011787 | 0.011989 | 0.009325629 | 0.023247895 |
| K08978 | bacterial/archaeal transporter family protein | 1.899764 | 0.008234 | 0.008326 | 0.009106418 | 0.024784274 |
| K00651 | homoserine O-succinyltransferase/O-acetyltransferase | 1.881026 | 0.002953 | 0.002963 | 0.008456631 | 0.023464036 |
| K20861 | FMN hydrolase / 5-amino-6-(5-phospho-D-ribitylamino)uracil phosphatase | 2.063187 | 0.011787 | 0.011993 | 0.007491599 | 0.030423796 |
| K04027 | ethanolamine utilization protein EutM | 1.81211 | 0.002579 | 0.002588 | 0.002144374 | 0.014920324 |
